# Distribution and Temporal Changes of Autoantibody-Mediated Pathogenic Mechanisms Among Acetylcholine Receptor-Positive Myasthenia Gravis Patients

**DOI:** 10.1101/2024.08.28.24312367

**Authors:** Fatemeh Khani-Habibabadi, Bhaskar Roy, Minh C Pham, Abeer H Obaid, Beata Filipek, Richard J Nowak, Kevin C O’Connor

## Abstract

**Objective:** Given that acetylcholine receptor-positive (AChR+) myasthenia gravis (MG) is mediated by AChR- specific autoantibodies, the emergence of new therapeutics underscores the importance of investigating AChR-specific autoantibody repertoire. This study aimed to assess the distribution of AChR-specific autoantibody isotypes, IgG subclasses, and the pathogenic mechanisms they mediate in AChR+ MG patients. Furthermore, we investigated longitudinal changes in autoantibody repertoire and the associated pathogenic mechanisms.

**Methods:** Serum samples (N=210) from 50 AChR+ generalized MG patients collected longitudinally over two years as part of the B-Cell Targeted Treatment in MG (BeatMG) study were evaluated using a set of cell-based assays.

**Results:** In cross-sectional samples, IgA and IgM AChR-specific autoantibodies were observed in the co-occurrence of IgG in 10% and 12% of patients, respectively. Among them, 4% had all three isotypes. AChR-IgG1 was found in 67.4%, followed by IgG3 (21.7%) and IgG2 (17.4%). Complement was active in 84.8%, followed by AChR internalization (63%) and blocking (30.4%). Complement and AChR internalization were simultaneously active in 45.6%, complement and blocking were active in 10.8%, and all three pathomechanisms were active in 17.4%. Blocking alone was active in only 2.1%; AChR internalization alone was not found. Temporal fluctuations of autoantibody isotypes/ IgG subclasses and the associated pathogenic mechanisms were observed.

**Interpretation:** These results demonstrate that a subset of patients have autoantibodies that can mediate pathogenic mechanisms and include isotypes/IgG subclasses that current therapeutics may not effectively target. Accordingly, defining individual patient AChR-specific autoantibody profiles may afford more accurate application of therapeutics designed to target specific autoantibody-mediated mechanisms.

## Introduction

Acetylcholine receptor (AChR)-specific autoantibodies, present in many patients with myasthenia gravis (MG), disrupt neuromuscular signal transmission by mediating three distinct pathogenic mechanisms^1–7^: (i) complement activation, (ii) AChR crosslinking, stimulating receptor internalization and degradation (known as antigenic modulation), and (iii) acetylcholine (ACh) binding site blocking (receptor antagonism). Most AChR-specific autoantibodies are of the IgG isotype (AChR-IgG) and can include different IgG subclasses^8–12^. AChR-IgM and IgA are also present alongside IgG, although they are less common and typically found at lower titers^13, 14^. Individual human AChR-specific monoclonal autoantibodies can mediate one, two, or three pathogenic mechanisms with varying efficiency^6, 7^. These efficiencies are partly attributed to the AChR subunits and epitopes they bind to, the strength of binding, and cooperative interactions^6, 7^. Additionally, the AChR-specific autoantibody isotypes and IgG subclasses play a significant role in pathology, especially in the case of complement activation. Serum AChR-specific autoantibodies are polyclonal, encompassing a heterogeneous pool of subunit and epitope specificities, isotypes and subclasses. Consequently, they mediate the pathogenic mechanisms through a variety of AChR-specific clones that can differ considerably between patients.

The emergence of therapeutics targeting AChR-specific autoantibodies has highlighted the importance of understanding the heterogeneity of the AChR-specific autoantibody repertoire. These therapeutics include inhibitors of the complement pathway (eculizumab, ravulizumab, and zilucoplan) that interrupt the formation of the membrane attack complex (MAC)^15–17^. Additionally, the neonatal Fc receptor (FcRn) inhibitors, efgartigimod^18^ and rozanolixizumab^19^, reduce circulating IgG levels, including AChR-specific autoantibodies. While both therapeutic approaches can lead to considerable clinical improvement in MG patients, a subset fails to respond. The variability in therapeutic response may be partly attributed to heterogeneity in AChR-specific autoantibodies repertoire. While complement inhibitors may be effective in some individuals, the contribution of autoantibody-mediated AChR internalization and blocking may continue to manifest pathology in others. Similarly, inhibiting FcRn-mediated IgG recycling can yield desired clinical outcomes by lowering AChR-IgG, but the presence of AChR-IgM or IgA isotype, which FcRn does not recycle, may contribute to poorer outcomes.

These collective laboratory and clinical findings have exposed our incomplete understanding of the characteristics of the circulating autoantibody repertoire in AChR+ MG patient populations. Specifically, we need to further investigate isotype/IgG subclass usage, the representation of each pathogenic mechanism, and whether this repertoire can change over time. To this end, we leveraged a cohort of MG patient-derived serum specimens collected over two years at four additional 6-month separated timepoints. Using a suite of live cell-based assays, we measured AChR-specific autoantibody isotypes, IgG subclasses, and the efficiency of associated pathogenic mechanisms in these specimens.

## Methods

### Human serum samples

This study was approved by the Yale University Institutional Review Board. A centralized institutional review board and independent ethics committee approved the study protocol and all amendments. Informed written consent, according to the Declaration of Helsinki, was received from all participants prior to their inclusion in this study. Serum samples were sourced from 50 AChR+ gMG patients with extensively documented clinical profiles obtained longitudinally as part of the BeatMG study (<u>clinicaltrials.gov</u>: NCT02110706)^20^ over two years, and timepoints separated by 6 months (**Supplementary Fig. 1 and Supplementary Table 1**).

### Measurement of autoantibody-mediated pathogenic mechanisms

Serum AChR-specific autoantibody capacity to activate complement was assessed by measuring MAC deposition on AChR-expressing triple-knockout HEK293T cells devoid of complement regulator genes CD46, CD55, CD59 as well as by measuring C3d deposition on AChR-expressing HEK293T cells, as previously described^21, 22^. The capacity of serum autoantibodies to block α-bungarotoxin (α-BTX, serving as a proxy for ACh) binding to AChR was evaluated using AChR-expressing HEK293T cells, as previously described^6, 23^. Autoantibody-mediated AChR internalization was assessed using the human rhabdomyosarcoma CN21 muscle-type cell line^24^, as previously described^25^. AChR-specific autoantibody capacity to internalize surface AChR was alternatively assessed using the Zenon™ pHrodo™ iFL IgG Labeling Reagent (ThermoFisher, Z25612) to label the IgG Fc region, which subsequently became fluorescent and visualized in the acidic milieu of the endosome. Extended detailed methods are available in the **Supplementary Methods**.

## Results

### Experimental design

This investigation aimed to define the circulating AChR autoantibody repertoire in MG patients and describe how the three different pathogenic mechanisms are represented both between and within patients over time. The serum samples (N=210) were rigorously collected longitudinally over two years from 50 AChR+ MG patients as a part of a B cell depletion (utilizing rituximab) clinical trial at 5 timepoints (including baseline) separated by 6 months. The placebo and treatment groups included 26 and 24 participants, respectively (**Supplementary Table 1 and Supplementary Fig. 1**). Baseline and placebo group specimens were used for investigating the repertoire both cross-sectionally and longitudinally. The study design has three sections. First, using baseline samples (N=50) autoantibody isotype/subclass distribution, the associated pathogenic mechanisms and associations between the assay-derived data and clinical metrics were investigated. Second, a longitudinal analysis on serum samples collected from the placebo group (N=26) was performed to observe how autoantibody isotype, subclass, and pathogenic mechanisms change over time. Third, the influence of B cell depletion on the autoantibody repertoire and the associated pathogenic mechanisms was investigated. We used established live rapsyn-clustered AChR cell-based assays which we developed to quantitatively measure the distribution of autoantibody isotypes (IgG, IgM, and IgA) and IgG subclasses (IgG1, IgG2, IgG3, and IgG4). Additional modifications were made to adapt it for measurement of AChR autoantibody-mediated pathogenic mechanisms, including classical complement pathway activation, AChR internalization, and blocking of the ACh binding site.

### Autoantibody isotype and IgG subclass distribution

The autoantibody isotypes (IgG, IgM, and IgA) and four IgG subclasses were measured in the baseline serum samples (N=50) from both rituximab and placebo groups using a set of CBAs. Recombinant human AChR mAbs with IgG subclass-specific Fc were produced by subcloning the variable region of the AChR mAb-637 IgG1^26^ and mAb-03 IgG1^6^ into IgG2, IgG3, and IgG4 subclass expression plasmids for using as assays positive controls. Recombinant mAbs, with isotype-specific Fc, unambiguously demonstrated that the commercial secondary antibodies used in the CBAs are highly specific for the isotypes and IgG subclasses (**Supplementary Fig. 2 and 3**). AChR-IgG binding capacity was observed in 46 out of 50 samples. The four negative samples in the binding assay had very low titer by RIA and were below the assay sensitivity (**Fig. 1A**). IgA (5/50; 10 %) and IgM (6/50; 12%) isotypes were also observed, however less frequently, and always in co-occurrence with IgG (**Fig. 1, B and C**). The IgG subclasses were measured in the 46 samples that showed AChR-IgG binding. The IgG1 subclass was observed in 31/46 (67.4%), followed by lower frequencies of IgG2 (8/46, 17.4%) and IgG3 (10/46, 21.7%; **Fig. 1D-F**). AChR-IgG4 was not detected in any sample (**Fig. 1G**). Irrespective of isotype or IgG subclass representation, the binding capacity varied considerably among the tested samples.

**Figure 1.**
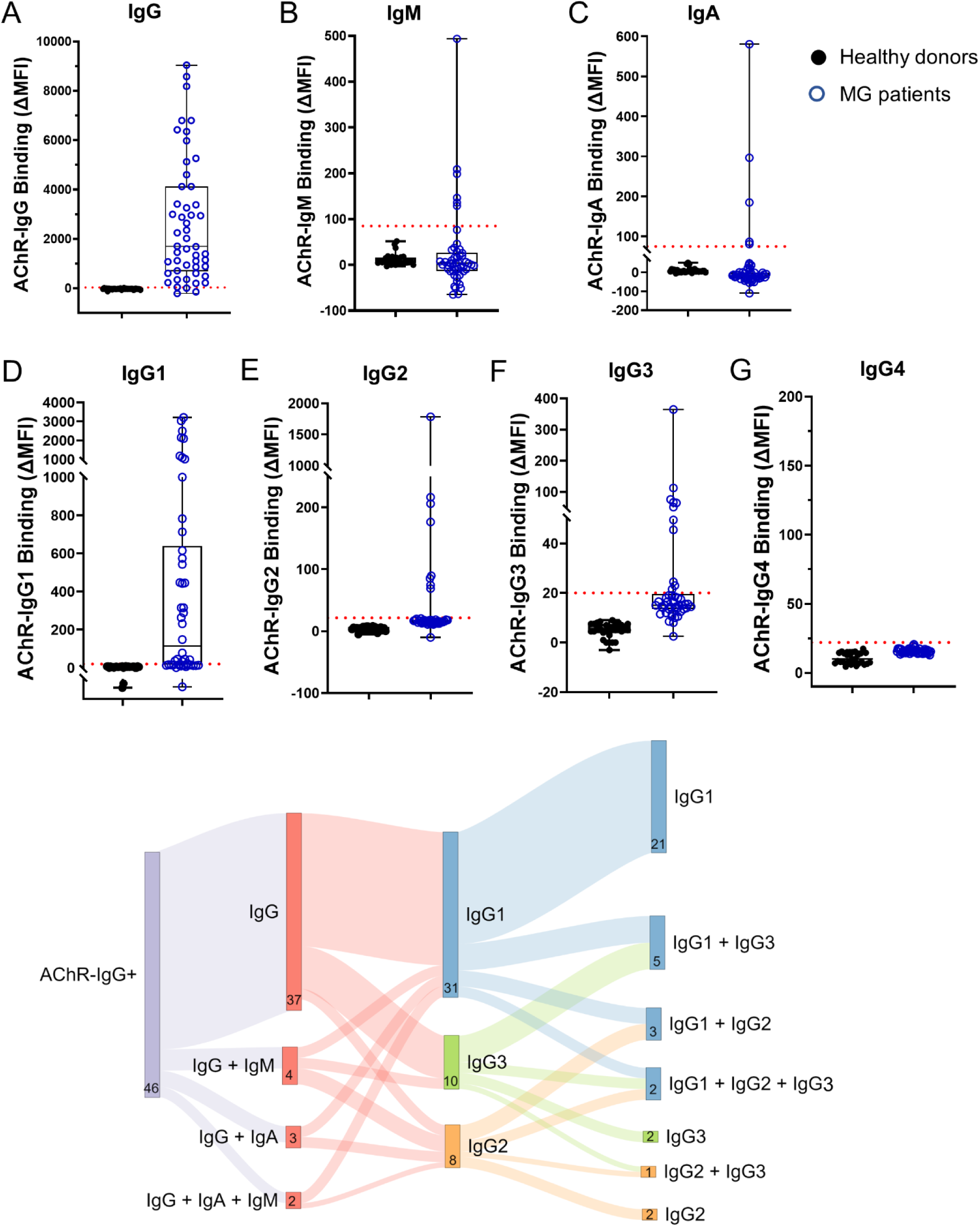
Distribution of AChR-specific autoantibody isotypes and IgG subclasses. Serum samples collected from AChR+ MG patients at baseline (N=50) were tested for the presence of AChR autoantibody isotypes and IgG subclasses using AChR-specific cell-based autoantibody binding assays. The binding capacity of AChR-specific (**A**) IgG, (**B**) IgM, and (**C**) IgA is shown. Patients with AChR-IgG (N=46) were further examined for AChR-IgG subclasses: (**D**) AChR-specific IgG1, (**E**) IgG2, (**F**) IgG3, and (**G**) IgG4. (**H**) Sankey diagram showing the overall distribution of AChR-specific isotypes and IgG subclasses in patients with AChR-IgG (N=46). Each data point shown (**A-G**) represents the mean of experimental triplicates. The dotted line represents the antibody detection threshold (HD mean + 3×SD).

We next examined the co-occurrence of isotypes and IgG subclasses. AChR-IgG alone was observed in 37/50 (74%). AChR-IgG and IgA were observed in 3/50 (6%), and IgG and IgM together in 4/50 (8%). Co-occurrence of all three IgG, IgA, and IgM AChR autoantibodies were observed in 2/50 (4%). Neither IgM nor IgA were found without IgG. The IgG subclass distribution was represented principally by IgG1; 21/46 (45.6%) individuals harbored only IgG1, while 2/46 (4.3%) patients had IgG2, and 2/46 (4.3%) had IgG3 alone. The co-occurrence of IgG1 and IgG2 was observed in 3/46 (6.5%) individuals, and IgG1 co-occurred with IgG3 in 5/46 (10.9%). One individual had only IgG2 and IgG3 (2.2%). The co-occurrence of IgG1, IgG2, and IgG3 was detected in two samples (4.3%, **Fig. 1H**). Taken together, these data demonstrate that there are different distributions and combinations of AChR autoantibody isotypes and IgG subclasses among MG patients.

### Complement activation is the predominant pathogenic mechanism mediated by AChR-specific autoantibodies

The distribution of AChR autoantibody-mediated pathogenic mechanisms was investigated in baseline samples with detectable AChR-specific autoantibody binding (N=46). To evaluate the complement activity, we measured MAC deposition, the terminal product of the complement pathway. The complement assay showed that 35/46 (76%) of serum samples with AChR-IgG binding capacity had autoantibodies capable of activating the complement pathway as measured by MAC deposition on the cell surface (**Fig. 2A**). Since MAC forms pores in the cell membrane that lead to cell death, some cells may be lysed before FACS analysis, thereby underestimating the efficiency of serum samples in activating complement. To address this issue, we developed a complementary complement assay to detect an earlier product of the complement pathway: C3d, a membrane-bound complement component. We used C8- depleted serum to prevent complete MAC formation and subsequent cell lysis. In close agreement with the detection of complement activity by MAC, 39 out of 46 serum samples with AChR-IgG binding capacity (84.8%) showed C3d deposition, inclusive of the 35 detected by MAC deposition (**Fig. 2B**).

**Figure 2.**
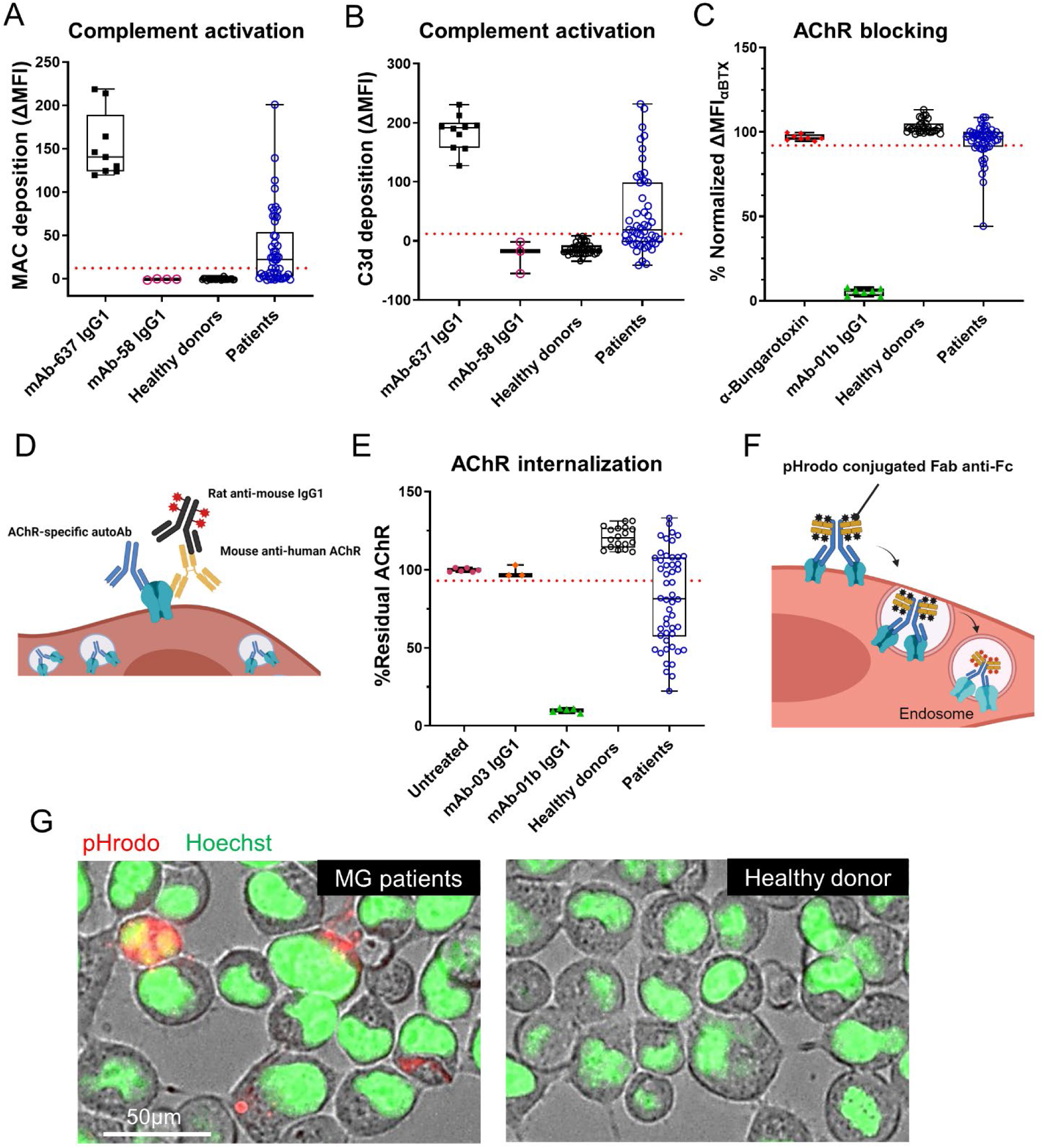
Measurement of AChR autoantibody-mediated pathogenic mechanisms. AChR IgG-positive serum samples collected at baseline (N=46) were examined to measure AChR autoantibody-mediated pathogenic mechanisms, including complement activation, receptor internalization (modulation), and ACh binding site blocking (receptor antagonism), using cell-based assays. The capacity of serum antibodies to activate complement was measured by (**A**) MAC and (**B**) C3d deposition on the surface of AChR-expressing HEK293T cells. The capacity of serum AChR-specific autoantibodies to block the ACh binding site (**C**) was measured using MFI values that were normalized to the untreated control (no mAb) as the upper limit (set to 100%). Schematic diagram, (**D**) illustrating the detection of surface AChRs using mouse anti-human mcAb-3 in the AChR internalization assay. The capacity of AChR-specific autoantibodies to internalize AChR (**E**) on the surface of CN21 cells. MFI values were normalized to the untreated control (no mAb) as the upper limit (set to 100%). Schematic diagram (**F**) illustrating the mechanism of pH-sensitive Zenon™ pHrodo™ iFL IgG labeling reagent used to visualize the internalization of AChR-specific IgG following binding and crosslinking surface AChRs. Images (**G**) of internalized IgG bound to AChR using fluorescence microscopy. Each data point (**A-C**, **E**) represents a mean of experimental triplicates. The dotted lines represent the detection threshold (HD mean + 3×SD for the complement assay and HD mean −3×SD for the blocking and internalization assays).

We next evaluated the capacity of AChR-specific autoantibodies to block the ACh binding site using α-BTX. In this assay, blocking autoantibodies occupy the ACh binding site and interfere with the fluorescent-labeled α-BTX binding to AChR, thereby causing a measurable decrease in the α-BTX fluorescent signal. Blocking assay demonstrated that 14/46 (30.4%) of serum samples with AChR-IgG binding capacity had blocking autoantibodies (**Fig. 2C**).

The ability of serum autoantibodies to internalize AChR was assessed by detecting residual AChR on the cell surface. First, residual AChR on the cell surface was measured utilizing mouse anti-human mcAb-3^27^ (**Fig. 2D** and **Supplementary Fig. 4A**). We found that 29/46 (63%) of serum samples with AChR-IgG binding capacity included autoantibodies that internalize AChR (**Fig. 2E**). As a complementary approach, internalization of AChR-IgG was measured using Zenon™ pHrodo™ iFL IgG Labeling Reagent, a pH-sensitive dye (**Fig. 2F**). After exposure to the acidic milieu of the endosome, the pHrodo fluorescent signal was quantified using flow cytometry (**Supplementary Fig. 4B**) and visualized using fluorescence microscopy (**Fig. 2G** and **Supplementary Fig. 4C and D**). The magnitude of AChR internalization in a subset of samples was measured using both assay approaches, demonstrating that the two assays are comparable for measuring AChR internalization (**Supplementary Fig. 4E**). As observed with the AChR-specific autoantibody binding capacity, the magnitude of complement activation, AChR internalization, and blocking varied considerably among individuals.

We next examined the co-occurrence of the three autoantibody-mediated pathogenic mechanisms. Complement activation was the principal (76-84.8%) pathogenic mechanism mediated by autoantibodies. However, complement activity alone was observed in only 5/46 (10.8%) patients; rather, it was more often found in combination with AChR internalization (21/46; 45.6%), blocking (5/46; 10.8%), or simultaneous occurrence of all three pathogenic mechanisms (8/46; 17.4%). Only 1/46 (2.1%) exclusively carried blocking autoantibodies. Six out of 46 (13%) serum samples did not produce signals above background in all three pathogenic mechanism assays; these samples displayed a low binding capacity for AChR-IgG as well (**Fig. 3**). In summary, these data suggest that complement activation (84.8%) is the principal autoantibody-mediated pathogenic mechanism among patients, followed by AChR internalization (63%) and blocking (30.4%). Complement activity was mainly found in combination with both or either AChR internalization or blocking.

**Figure 3.**
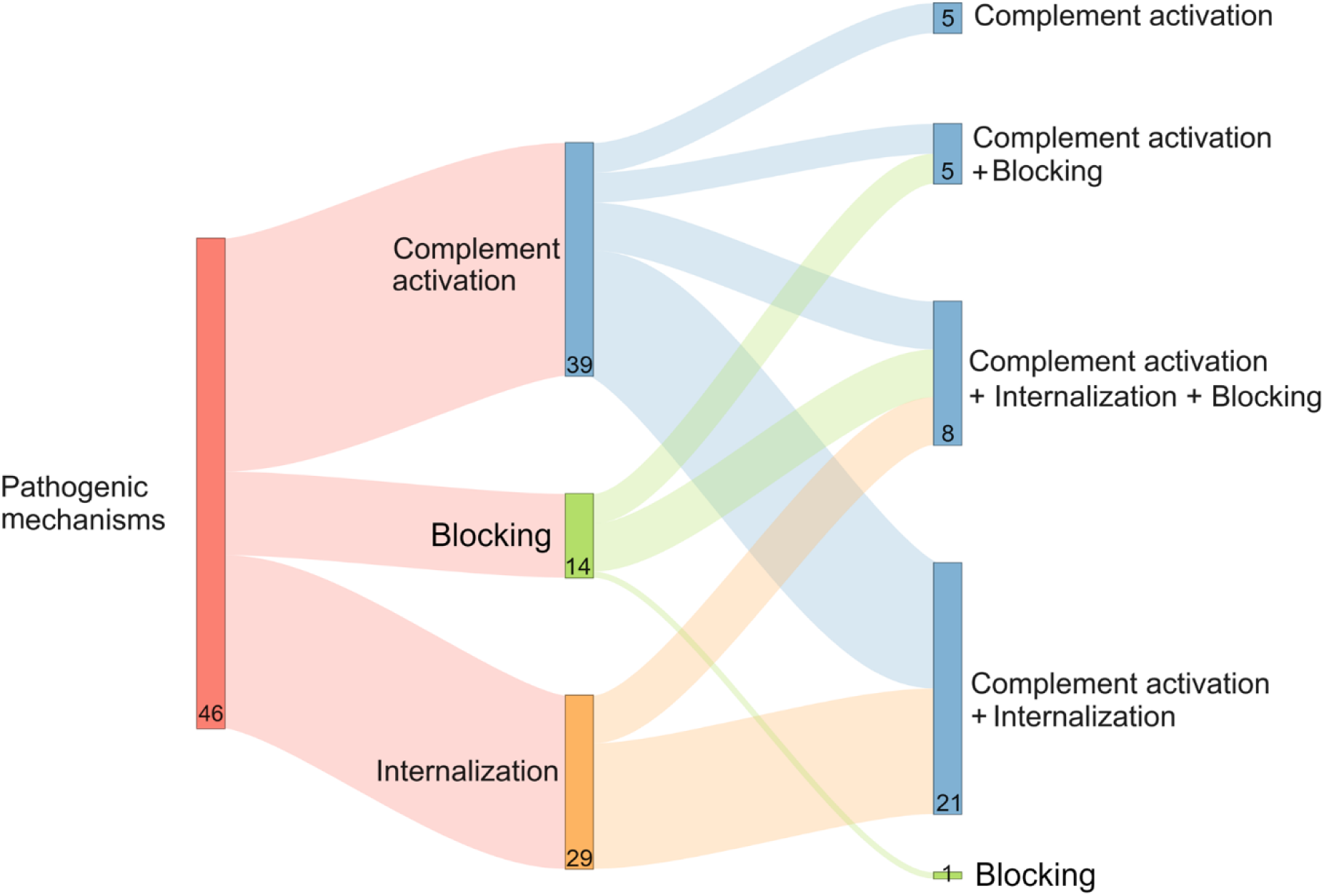
Distribution of AChR autoantibody-mediated pathogenic mechanisms. The Sankey diagram illustrates the distribution and overlap of AChR autoantibody-mediated pathogenic mechanisms at the baseline timepoint of the patient cohort (N=46).

### AChR-IgG binding capacity is associated with complement activity and AChR internalization

Association between the autoantibody binding capacity and the magnitude of the associated pathogenic mechanisms was investigated. Samples with stronger AChR-IgG binding capacity most often activated complement and internalized AChR with higher efficiency, suggesting a positive association between binding capacity and the effectiveness of the pathogenic mechanisms. However, the magnitude of blocking was not associated with binding capacity (**Fig. 4A**). Next, the correlation between the isotype/IgG subclass binding capacity and the associated pathogenic mechanisms was investigated in individuals who showed AChR-IgG binding in CBA (N=46). Correlation analysis demonstrated a significant association between the binding of AChR-IgG and MAC (r= 0.82, P<0.0001) and C3d (r= 0.69, P<0.0001) deposition, as well as AChR internalization (r= 0.85, P<0.0001). MAC and C3d deposition were also found to correlate with one another (r= 0.87, P<0.0001) and with the magnitude of AChR internalization (r= 0.84, P<0.0001; r= 0.74, P<0.0001; respectively). However, we did not observe an association between isotype/IgG subclass binding, complement activity, and AChR internalization with blocking. MAC deposition correlated with IgG1 (r= 0.35, P= 0.017), IgG2 (r= 0.34, P= 0.019), and IgG3 (r=0.47, P= 0.001), and C3d deposition was associated with IgG1 (r= 0.36, P= 0.013) and IgG3 (r= 0.44, P= 0.002, **Fig. 4B**). Quantitative myasthenia gravis (QMG), myasthenia gravis activities of daily living (MG-ADL), and myasthenia gravis composite (MGC) scores did not show any significant correlations with the isotype/IgG subclass binding capacity or their associated pathogenic mechanisms. Other clinical metrics, including disease duration, age of onset, and MGFA classification were not associated with either binding capacity or the magnitude of pathogenic mechanisms (**Supplementary Fig. 5**). Taken together, these data demonstrate that AChR-IgG binding capacity is associated with the magnitude of complement activity and AChR internalization but not with the blocking of ACh binding site.

**Figure 4.**
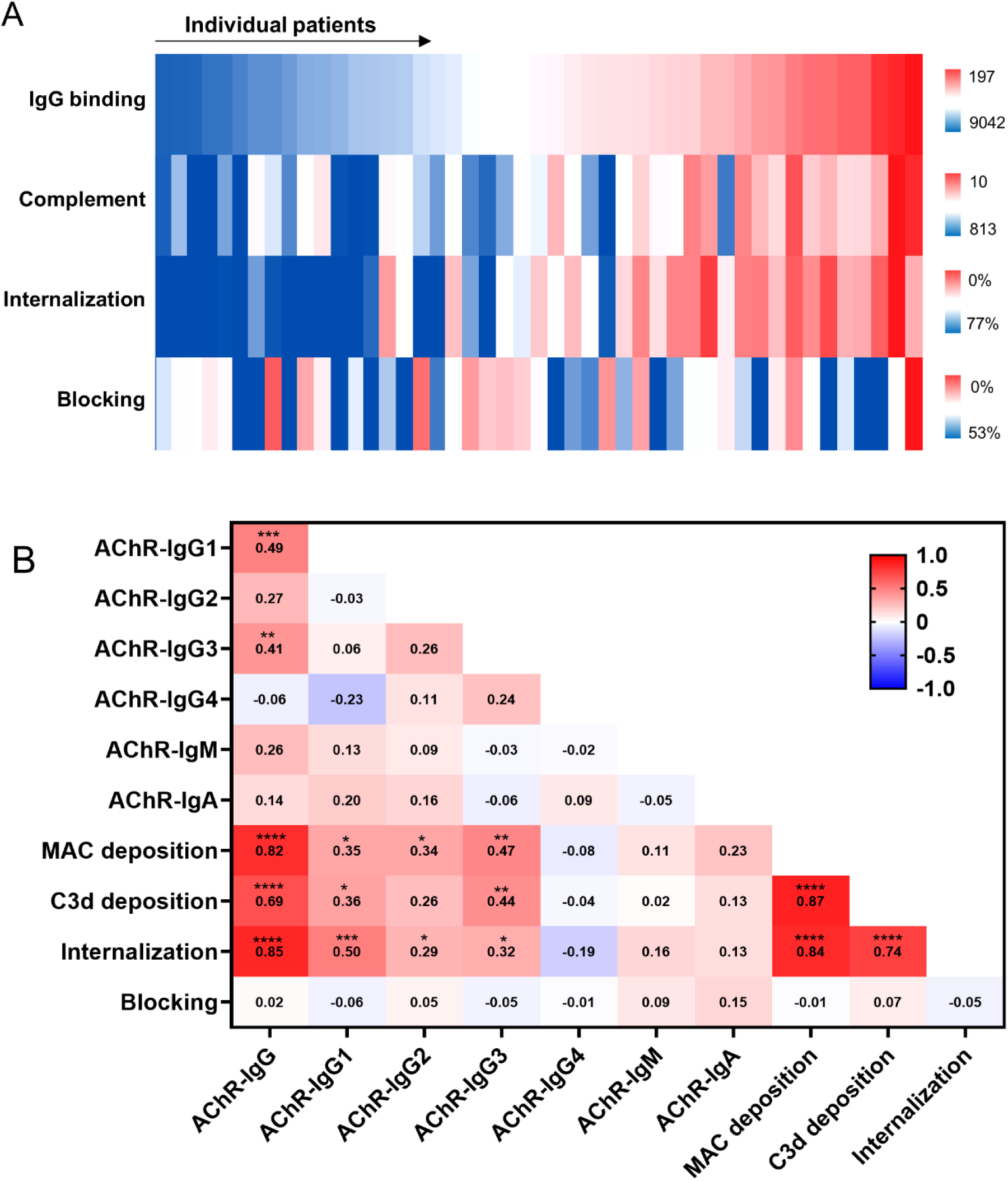
Association between autoantibody binding capacity and pathogenic mechanisms. Serum samples, collected at baseline, were examined to investigate how autoantibody binding capacity and autoantibody-mediated pathogenic mechanisms associate. Heatmap (**A**) showing the AChR-IgG binding capacity, and the magnitude of autoantibody-mediated pathogenic mechanisms measured in each individual (N=46). Each column of the heatmap represents the mean of triplicate tests for an individual patient. Each bar shows ΔMFI values for binding capacity, complement activity, or the percentage for AChR internalization and blocking. Scales are shown to the right of each column. Heatmap (**B**) showing the Spearman correlation analysis, calculated using ΔMFI values for each individual patient, between AChR- specific autoantibody binding and the associated pathogenic mechanisms at the baseline timepoint for the AChR-IgG positive patient cohort (N=46). The correlation coefficients (r) are shown on the heatmap. A significance threshold of P < 0.05 was used and is shown on plots when significance was reached: *P < 0.05; ** P < 0.01, *** P < 0.001, and **** P < 0.0001.

### AChR-IgM activates complement most efficiently

Considering the positive and robust association among AChR-specific autoantibody binding capacity, complement activation and AChR internalization, we further investigated the differential ability of IgA, IgM, and IgG subclasses in mediating these pathogenic mechanisms. To this end, we produced recombinant AChR mAbs with IgA, IgM, and IgG subclass-specific Fc by subcloning the variable region of the AChR mAb-637 IgG1 into IgA, IgM, and IgG2, IgG3, and IgG4 subclass expression plasmids. The IgM plasmid was co-transfected with the J chain to express IgM in its pentameric form, the predominant structure found in the serum^28^. Additionally, recombinant AChR mAb-03 IgG subclasses were produced by subcloning the variable region of the mAb-03 IgG1 into IgG2, IgG3, and IgG4 subclass expression plasmids. We expressed, quantified, and validated isotype and IgG subclass specificity and their binding to AChR (**Supplementary Fig. 2 and 3**). When we performed the binding assay with serial dilutions of the mAbs, all four subclasses of mAb-637 and mAb-03 showed similar binding to AChR. Similarly, mAb-637 IgA and IgM showed efficient binding to AChR. A human monoclonal derived from a neuromyelitis optica patient, specific to AQP4 (mAb-58 IgG1)^29^ was used as a negative control (**Fig. 5A-C** and **Supplementary Fig. 6A**). AChR-IgM was the most efficient at activating complement compared to other isotypes. AChR-IgG3 was the second most efficient complement activator, followed by IgG1. In contrast, IgA, IgG2, and IgG4 were inefficient at activating complement, while AQP4-IgG1 did not activate complement on AChR-expressing cells, as expected (**Fig. 5D** and **Supplementary Fig. 6B**). Next, we tested the mAb-637 isotypes and IgG subclasses in the internalization assay. Our data suggests that IgM is the most efficient at AChR internalization, followed by IgA and the four IgG subclasses (**Fig. 5E**). These data indicate that AChR-IgM is the most efficient at complement activation and AChR internalization. AChR-IgG3 is the most efficient complement activator amongst the IgG subclasses. AChR-IgA is inefficient in activating complement but is effective at AChR internalization.

**Figure 5.**
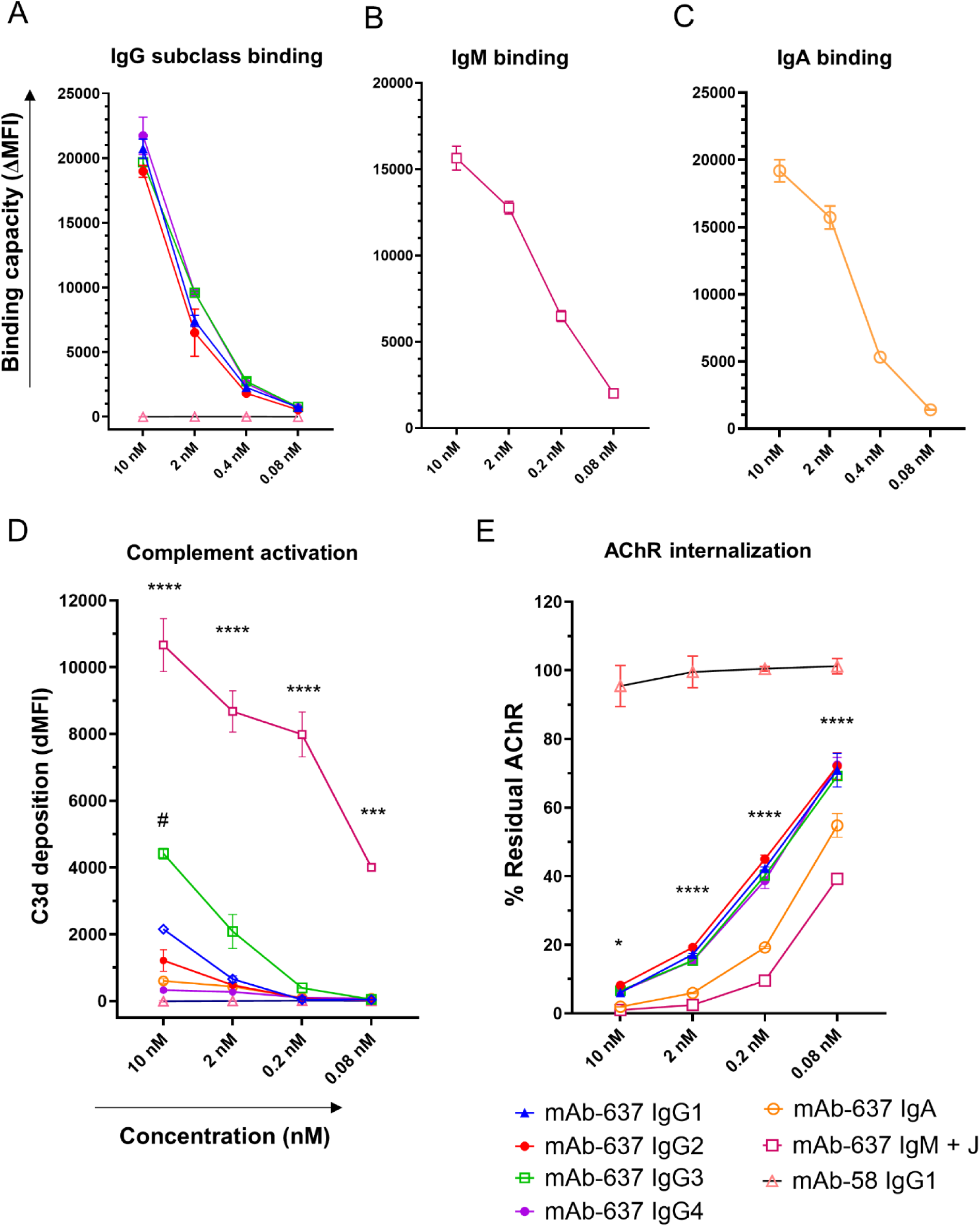
The efficiency of AChR-specific autoantibody isotype and IgG subclasses in activating complement and mediating AChR internalization. The variable region heavy chain of AChR mAb-637 was subcloned into antibody isotype and IgG subclass-expression vectors to assess differences in their ability to bind, activate complement, and internalize AChR. Titration plots show binding for mAb-637 (**A**) IgG subclasses, (**B**) IgM, and (**C**) IgA tested over a range of concentrations. Titration plot curves for (**D**) C3d deposition and (**E**) AChR internalization are shown for mAb-637 IgG subclasses, IgA, and IgM tested over a range of concentrations. The mAb-58 IgG1(AQP4-specific) was included as a negative control. Each data point (**A-C**, **E**) represents a mean of experimental triplicates, and error bars represent mean ± SD. Two-way ANOVA was used, followed by Tukey’s post-test. A significance threshold of P < 0.05 was used and is shown on plots when significance was reached: *P < 0.05; ** P < 0.01, *** P < 0.001, and **** P < 0.0001. Asterisks (*) compare mAb-637 IgM to IgG3; octothorpes (#) compare IgG3 to IgG1.

### The circulating repertoire of AChR-specific autoantibodies is variable during the disease course for individual patients

Considering the heterogeneity among individuals, we next examined patient samples that were collected longitudinally (every six months over two years) from the placebo group (N=26 patients, N=128 samples) to investigate how the repertoire of AChR-specific autoantibodies changes over time in individual patients. The longitudinal analysis of IgG binding and associated pathogenic mechanisms included serum samples below the detectable threshold in the IgG binding assay (baseline samples; N=26) to investigate potential changes during the disease course. Additionally, a subset of 13 patients were selected to be examined for longitudinal changes in AChR-IgM, IgA, and IgG subclasses. This analysis showed that the temporal binding capacity of AChR-IgG (N=26, **Fig. 6A**), IgA, IgM, and IgG subclasses underwent significant fluctuations (N=13; **Supplementary Fig. 7A-F**). Similarly, the magnitude of complement activity (C3d deposition), AChR internalization, and blocking underwent observable fluctuations within each individual over time, which were variable from one patient to another (N=26, **Fig. 6B-D**). Five representative patients were selected to show the AChR-specific autoantibody temporal changes in more detail. The combination of AChR-specific isotypes/IgG subclasses, the associated pathogenic mechanisms, and their fluctuation trend varied among patients. For example, patients 2 and 4 showed a sharp decrease and increase in IgG1 binding along with the associated pathogenic mechanisms, respectively. Patient 3 showed a high IgA binding capacity, while patient 5 showed a change in IgA binding capacity and the emergence of IgG1 and IgG3 by week 96, which corresponded with an increase in complement activity. The change in AChR-specific autoantibody binding was associated with the change in pathogenic mechanisms; however, disease severity was not associated with autoantibody binding or the associated pathogenic mechanisms (**Fig. 6E**).

**Figure 6.**
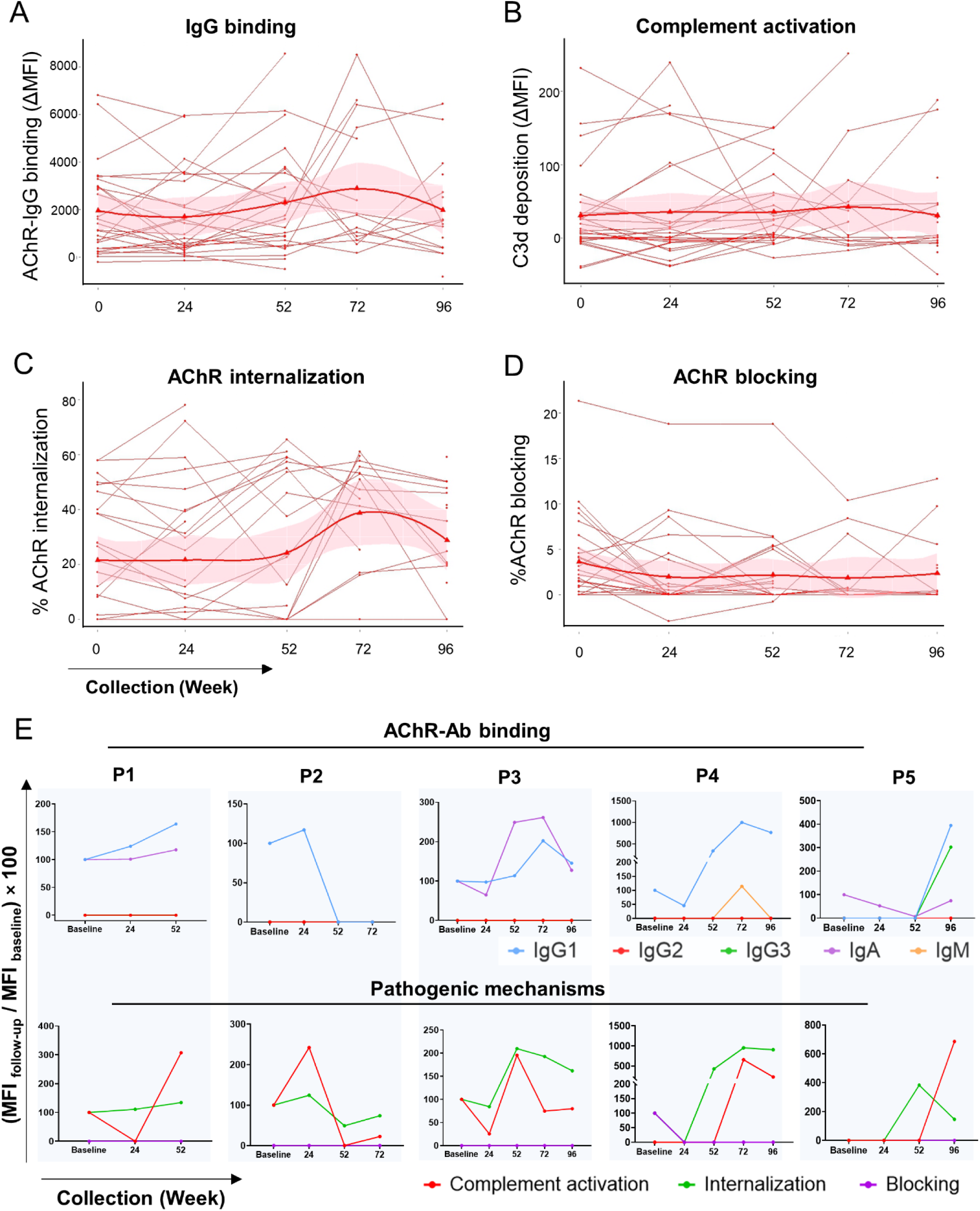
Temporal changes in autoantibody binding capacity and the magnitude of antibody-mediated pathogenic mechanisms. Serum samples collected longitudinally from placebo group (N=26) were examined to determine how the capacity of AChR autoantibody binding and the associated pathogenic mechanisms changed within each patient over two years. Spaghetti plots show temporal changes in (**A**) AChR-IgG binding capacity and the magnitude of (**B**) C3d deposition, (**C**) AChR internalization, and (**D**) blocking. Each line represents a single patient (mean of experimental triplicates). Solid lines connect collection timepoints. The trend line (smooth bold curve) was created using the locally estimated scatterplot smoothing (LOESS) method. The shaded area on the graph depicts a 95% confidence interval (CI). The AChR internalization and blocking percentages were calculated as (100 - (% residual AChR or % reduction of αBTX MFI)). Spaghetti plots (**E**) showing temporal changes in the frequency of AChR-specific isotypes and IgG subclasses and the associated pathogenic mechanisms in five representative patients in the placebo group. Each column displays an individual patient; the top row shows AChR-specific autoantibody isotype and IgG subclass binding and the associated pathogenic mechanisms in the bottom row. The change in binding capacity and pathogenic mechanisms is shown as a percentage change from baseline, calculated as (MFI follow-up / MFI baseline) × 100. If the baseline value was 0, the positive detection threshold (mean ΔMFI + 3×SD of healthy donors) was used instead.

Similar to the placebo group, patients in the rituximab group (N=24) showed a wide range of temporal changes over time. Because six patients from both rituximab and placebo groups discontinued the trial after week 52 (**Supplementary Fig. 1**), statistical tests were conducted on data from baseline, week 24, and week 52 to maintain analytical rigor. While 16/24 (66.7%) patients showed reduced AChR-IgG binding in response to rituximab by week 52, longitudinal analyses showed that rituximab treatment had minimal effects on the associated pathogenic mechanisms. Longitudinal statistical analysis comparing changes between the rituximab (N=24) and the placebo (N=26) showed that AChR-IgG binding did not change at week 24 compared to baseline; however, at week 52, a decrease in AChR-IgG binding capacity was observed in the rituximab group (p=0.004; **Supplementary Fig. 8A**). C3d deposition and AChR internalization did not show a significant change (**Supplementary Fig. 8, B and C**), but AChR blocking decreased at week 24 (p=0.016) and week 52 (p=0.027; **Supplementary Fig. 8D**). Additionally, in a subset of patients (N=10 rituximab, N=13 placebo) AChR-IgA binding did not change over time (**Supplementary Fig. 9A**). AChR-IgM binding decreased at week 24 (p=0.033; **Supplementary Fig. 9B**). AChR-IgG1 binding capacity decreased only at week 52 (p=0.025), while the binding capacity of AChR-IgG2 and IgG3 remained unchanged (**Supplementary Fig. 9, C-E**). These data indicate that rituximab does not effectively change the AChR-specific autoantibody repertoire and the associated pathogenic mechanisms.

## Discussion

Understanding the distribution of AChR-specific autoantibody isotypes/IgG subclasses and the pathogenic mechanisms they mediate has become more critical, given the development of new therapeutic approaches for treating MG, which target specific pathogenic mechanisms or antibody isotypes. Thus, understanding the heterogeneity of these key disease characteristics in individual patients can inform therapeutic choices and deepen the understanding of clinical responses to therapeutic intervention, which are essential for personalized medicine. To this end, we conducted a longitudinal examination of MG patient-derived serum using live CBAs to quantify AChR-specific autoantibody isotypes and IgG subclasses, and measure the pathogenic mechanisms they facilitate, including autoantibody-mediated complement activation, AChR internalization, and blocking.

We chose to use live CBAs as this approach can be more sensitive than the radioimmunoassay (RIA)^30, 31^, presents the AChR in its native conformation at a density similar to the NMJ, and does not detect autoantibody binding to intramembrane and intracellular epitopes, which are unlikely to be pathogenic^32^. For complement and blocking assays, we leveraged clustered AChR as it offers higher sensitivity than unclustered AChR^33^. Regarding the internalization assay, we employed mcAb-3 to measure the residual AChR on the cell surface. The mcAb-3 binds to an extracellular epitope outside the ACh binding site, thereby not confounding the detection of internalization-inducing autoantibodies with those that block^27^, an issue that can occur through the use of bungarotoxin in internalization assay. In addition, we used a limited-clustered AChR-expressing cell line (CN21) as AChR clustering inhibits receptor internalization^34^, and transfection damages the lipidic membrane and interrupts receptor internalization. These optimizations were crucial for ensuring the accuracy and reliability of our assays.

We experimentally confirmed the specificity of secondary antibodies to ensure the accuracy of the isotyping assays. This was particularly critical given the sequence homology (approximately 90%)^35^, among human IgG subclasses that can lead to cross-reactivity of commercial antibodies, resulting in inaccurate measurements. However, the secondary antibodies presented some limitations. The polyclonal secondary antibody used for detecting IgG (all subclasses) was more sensitive than the IgG subclass-specific monoclonal secondary antibodies. This could have led to a slight underestimation in subclassing results, as ten serum samples with detectable AChR-specific IgG binding fell below the detectable threshold when quantified using IgG subclass-specific secondary antibodies.

The binding assay demonstrated that 46/50 patients had detectable AChR-IgG at the baseline. This high frequency was expected, given that the trial inclusion criteria focused on AChR+ MG patients. RIA titers in this sample cohort raged from 0-193 nmol/L. All samples with titer > 0.1 nmol/L showed AChR-IgG binding in the CBA. The four samples that were negative in the CBA provided titer values < 0.1 nmol/L. Overall, the binding data was consistent between the two approaches.

While our findings demonstrated IgG1 predominancy, other studies have reported variable frequencies of IgG subclasses; some indicate a predominance of IgG1, 2, or 3 and some suggest the co-occurrence of IgG4 with other subclasses^8, 10, 11^. Given the concerns regarding the cross-reactivity of commercial antibodies^9^, the frequency of IgG2, 3, and 4 may have been overestimated in some of these studies. In this sample cohort, we showed complement activity in 84.7% of individuals, which aligns with our previous study, showing a frequency of 69%^21^. The slight differences are likely attributable to the different patient cohorts and inclusion criteria. This data also aligns well with early studies of muscle biopsy tissue reporting IgG, C3, and MAC at the NMJ found in 84-100% of patients^2, 36, 37^. The successful long-term use of complement inhibitors in treating gMG further highlights the prevalent contribution of complement activity to MG pathogenesis^15, 38^. Likewise, internalization-inducer autoantibodies were found to be active in 63% of individuals. Previous studies have reported higher frequencies of 80-90%, which may be overestimated due to other assay methods employed. These include using ^[125I]^α-BTX, which detects internalization and blocking confoundedly^39^, and using non-human skeletal muscle culture^40^. Less commonly, we found that 30% of patients had autoantibodies that mediate blocking using our optimized CBA; however, the previous reported frequencies have been highly incongruous ranging from 5-95%^5, 8, 9, 33, 39–43^. The high level of disagreement among studies is difficult to explain. It has been shown that AChR autoantibodies block the Ach binding site more efficiently on AChR extract from denervated muscles than those from normal human muscles. These disagreements may be due to the different sources and the quality of the AChR extracts^44^.

It is evident that, within an individual, the autoantibody repertoire is polyclonal. It is conceivable that individual autoantibody clones within the polyclonal serum contribute to each pathogenic mechanism individually, but we also know that a single autoantibody clone can mediate all three^6^. Moreover, distinct combinations of clones can cooperate to enhance effector functionality^6, 7^. In addition, our data showed co-occurrence of AChR-specific isotypes/IgG subclasses in individual patients, aligning with previous investigations^8, 10, 11, 13, 14^. Therefore, the autoantibody profile in each individual is a unique heterogeneous combination of autoantibody properties and autoantibody-mediated pathogenic mechanisms.

We showed that AChR-IgM mAb robustly activated complement, followed by IgG3 and IgG1. This aligns with previous studies showing that IgM^28^ and IgG3^45, 46^ are superior to IgG1 in activating complement. AChR-IgM-mediated receptor internalization is more efficient than that of other isotypes, this result may be due to a multivalent cross-linking between AChRs mediated by the pentameric IgM. Given the varying capacities of different isotypes and IgG subclasses in mediating pathogenic mechanisms, it is conceivable that the autoantibody repertoire can influence treatment effectiveness if the therapy does not address all the pathogenic mechanisms involved in an individual patient. Specifically, C5 inhibitors target only the complement pathway, so patients with highly effective internalization-inducing and blocking autoantibodies may not benefit completely from this treatment. Conversely, FcRn blockers inhibiting IgG recycling will theoretically affect all pathogenic mechanisms^15–19^; however, some patients may not experience full clinical benefits if their repertoire includes other isotypes. For instance, patients with pathogenic AChR-IgM may not respond well to FcRn blockers. This highlights the importance of leveraging the autoantibody repertoire for individualized precision medicine.

The opportunity to examine AChR autoantibody isotypes, IgG subclasses, and their efficiency in mediating different pathogenic mechanisms over a uniform period demonstrated considerable variation within individuals. These findings suggest that, in some patients, the circulating autoantibody repertoire is changeable and can vary within a relatively short period. The most likely source of circulating AChR-specific autoantibodies is long-lived plasma cells (LLPC) residing in the bone marrow and, in some patients, among the lymphocytic infiltrate in the thymus^47^. The larger circulating autoreactome in MG patients is thought to be different among individuals but quite stable, showing slight variation, within each person^48^. Thus, it is unclear why the circulating AChR-specific autoantibody repertoire changes over time. This may point to the turnover of the autoantibody-producing LLPC compartment^47^. Additionally, it is important to consider the influence of treatment. The individuals in the placebo group, received steroids during the trial period, which may alter expression of the circulating AChR autoantibody repertoire.

Our analysis did not reveal a significant correlation between the AChR autoantibody properties we measured and clinical characteristics. While the lack of associations between autoantibody titer and disease severity is often characteristic of MG^11^, some studies showed a correlation between a change in autoantibody titer and disease severity^49, 50^. Autoantibody titer has also been associated with treatment outcomes^51, 52^. It is possible that our longitudinal data did not produce such an association because the patient cohort had relatively mild disease upon enrollment, making changes less conspicuous. The lack of correlation may also be explained by several other factors. For example, circulating autoantibodies may not accurately represent autoantibodies actively mediating pathology at the NMJ. Epitope specificity may also be associated with disease severity, as suggested by the fact that alpha-binding mAbs are more efficient at complement activation^6^. In addition, the glycosylation of autoantibodies could impact antibody-mediated effector functions and may also be associated with disease severity^53^.

We observed considerable temporal changes in the AChR-specific autoantibody repertoire in individuals treated with rituximab. This heterogeneous nature of the autoantibody repertoire led to varied individual responses, making it challenging to conclude how rituximab specifically affects AChR autoantibodies. In the BeatMG study, rituximab reduced the autoantibody titer, but did not demonstrate a difference in steroid-sparing effect as compared to placebo in a cohort of patients with mild-moderate AChR+ MG. As a phase 2 trial it was primary designed to assess safety and to provide a go/no-go decision for a future efficacy phase 3 trial^20^. On the other hand, the RINOMAX trial, an efficacy study, demonstrated that a greater proportion of patients achieved minimal disease manifestation in the rituximab group as compared to the placebo group. The RINOMAX cohort is distinct from the BeatMG cohort in that it enrolled patients within 2 years of diagnosis^54^. Rituximab targets the CD20-positive precursor of plasma cells, not the CD20-negative cells, i.e., plasma cells or most plasmablasts. Therefore, the superior clinical response in new-onset MG patients may be due to early inhibition of long-lived plasma cell formation.

Our study underscores the complexity and heterogeneity of the AChR-specific autoantibody repertoire. It provides clinical insights by suggesting that the autoantibody repertoire may be associated with how patients respond to the therapeutics. These findings highlight the need for precision medicine in selecting treatments that can address the diverse and dynamic nature of MG autoantibody repertoire. This study paves the way for further investigation into the relationship between antibody pathogenicity and factors such as clonality, epitope specificity, and antibody glycosylation. We suggest incorporating these assays into MG clinical trials to explore potential associations with outcomes prior to and during therapeutic administration.

## Acknowledgments

The authors thank Dr. Sarosh R. Irani of the Mayo Clinic (Jacksonville, Florida, USA) and Dr. David R. Martinez of Yale School of Medicine (New Haven, Connecticut, USA) for providing the IgM and IgA expression vectors, respectively. The mcAb-3, was provided by Dr. Vanda A. Lennon and Dr. John R. Mills of Mayo Clinic (Rochester, Minnesota, USA). The C3d-specific mAb was provided by Dr. Joshua M. Thurman of University of Colorado (Anschutz Medical Campus, Aurora, Colorado, USA), Dr. Kelly Fahnoe, and Dr. Stefan Wawersik of Q32 Bio (Waltham, Massachusetts, USA). The AQP4-specific human mAb58 was generously provided by Dr. Jeffrey L. Bennett of the University of Colorado Anschutz Medical Campus (Aurora, CO, USA). The authors would like to thank the BeatMG study investigators, study teams, and patients that participated in the phase 2 trial.

## Author Contributions

FKH and KCO designed the study, interpreted the data, and wrote the manuscript. FKH designed and validated methods, performed experiments, acquired, and analyzed data. BR analyzed, interpreted, and assisted in writing. MCP, AHO, and BF performed experiments, acquired and analyzed data. RJN provided study samples and clinical data. Figures were produced with GraphPad Prism, SAS Analytics Software, R, FlowJo, BioRender, and SankeyMATIC by FKH and BR. All authors contributed to the editing and revising of the manuscript.

## Data availability

Anonymized data will be shared at request by qualified investigators following the execution of appropriate materials transfer agreements.

