## Supplementary for "Distribution and Temporal Changes of Autoantibody-Mediated Pathogenic Mechanisms Among Acetylcholine Receptor-Positive Myasthenia Gravis Patients"

**Running head: AChR Autoantibody-Mediated Pathogenic Mechanisms**

Fatemeh Khani-Habibabadi, PhD[1,2], Bhaskar Roy, MD[1], Minh C Pham, BS[2], Abeer H Obaid, PhD[1,2,3], Beata Filipek, MD[1,2,4], Richard J Nowak, MD[1], and Kevin C O'Connor, PhD[1,2]

1. Department of Neurology, Yale School of Medicine, New Haven, CT, 06511. USA
2. Department of Immunobiology, Yale School of Medicine, New Haven, CT, 06511, USA
3. Institute of Biomedical Studies, Baylor University, Waco, TX, 76706, USA
4. Department of Pharmaceutical Microbiology and Biochemistry, Medical University of Lodz, Lodz, Poland.

Corresponding author: Kevin C. O’Connor, PhD
Yale School of Medicine
Departments of Neurology & Immunobiology
Room 353L
300 George Street
New Haven, CT 06511


**List of supplementary materials**

**Supplementary Methods**


- Production of recombinant AChR-specific monoclonal antibodies (mAbs)
- Isotype and IgG-subclass specific ELISA
- AChR-specific autoantibody isotyping
- Complement assays
- ACh binding site blocking assay
- AChR internalization assay
- Statistical analysis
- Data availability


**Supplementary Figures**


- Supplementary Fig. 1
- Supplementary Fig. 2
- Supplementary Fig. 3
- Supplementary Fig. 4
- Supplementary Fig. 5
- Supplementary Fig. 6
- Supplementary Fig. 7
- Supplementary Fig. 8
- Supplementary Fig. 9


**Supplementary Tables**


- Supplementary Table 1


**References**

### Supplementary methods

#### Production of recombinant AChR-specific monoclonal antibodies (mAbs)

To express AChR-IgA and IgM mAbs, DNA fragments encoding the variable region heavy chain of mAb-637 were designed, synthesized (Twist Bioscience, USA) and cloned into human IgA1 and IgM expression plasmids, generously provided by Dr. David R. Martinez (Yale School of Medicine, CT, USA) and Dr. Sarosh R. Irani (Mayo Clinic, Jacksonville, Florida, USA), respectively.

To express AChR-IgG subclass mAbs, IgG2, IgG3, and IgG4 expression plasmids were engineered from our human IgG1 heavy chain expression plasmid[1] using published human constant regions (GenBank; AXN93670.2, AK097307.1, KJ901516.1) to replace the constant region heavy chain of IgG1. The variable region heavy chains of AChR mAb-637[2] and mAb-03[3] were subcloned into the IgG2, IgG3, and IgG4 expression plasmids. The sequence integrity of the engineered heavy chain expression plasmids was confirmed using Sanger sequencing of the insert and sequencing the entire plasmids with the Oxford Nanopore platform (Plasmidsaurus, USA).

Recombinant mAbs were then expressed by transfection of heavy and light chain plasmid pairs into HEK293T cells (ATCC, CRL-3216) in the presence of linear polyethylenimine (Polysciences, 23966) and Nutridoma-SP (Roche, 11011375001). The IgM plasmid was co-transfected with the J chain (generously provided by Dr. Sarosh R. Irani, Mayo Clinic, Jacksonville, Florida, USA) to express it in its pentameric form. IgG antibodies were then purified from antibody-rich supernatants, collected on day 5, using Protein G Sepharose beads (Cytiva, 17-0618-02)[4].

#### Isotype and IgG subclass-specific ELISA

To confirm the expression of AChR-IgM and IgA mAbs and to measure their concentration in the antibody-rich supernatants, ELISA plates were coated with goat anti-human IgA+IgG+IgM (Jackson ImmunoResearch, 109-005-064) through incubation for 16 hours at 4 °C. Serial dilutions of the Human IgM (ThermoFisher, 39-50620-65) and human IgA standards (Sigma-Aldrich, I4036) ranging from 0.01 to 1 µg/mL, were included in the assay alongside serial dilutions of the antibody-rich supernatants. The presence of AChR-IgA or IgM was detected using peroxidase-conjugated goat anti-human serum IgA-α-chain-specific (Jackson ImmunoResearch, 109-005-011) or goat anti-human IgM-Fc5µ (Jackson ImmunoResearch, 109-001-043), respectively, at a dilution of 1:20,000. Additionally, to confirm the expression of the AChR-IgG1 mAb, a serial dilution of AChR-IgG1 was included in the assay, which were subsequently detected using a peroxidase-conjugated goat anti-human IgG antibody (109-035-098, Jackson ImmunoResearch, 1:20,000).

To confirm the expression of AChR-IgG subclass mAbs, ELISA plates were coated with subclass-specific antibodies for IgG2 (05-3500, Thermo Fisher Scientific), IgG3 (05-3600, Thermo Fisher Scientific) or IgG4 (MA5-16716, Thermo Fisher Scientific) at a concentration of 5 µg/mL. The plates were then incubated with serial dilutions of the mAbs ranging from 0.037 to 10 µg/mL and were subsequently detected using a peroxidase-conjugated goat anti-human IgG antibody (109-035-098, Jackson ImmunoResearch, 1:20,000). A standard protocol for colorimetric development, detection, and analysis of ELISA was applied.

**AChR-specific autoantibody isotyping**

AChR-specific autoantibody binding was tested using a suite of rapsyn-clustered live cell-based assays (CBAs) as previously described[5-7]. Briefly, HEK293T cells (ATCC, CRL3216) were transiently transfected with adult AChR (2α, β, δ, and ε) and rapsyn-GFP plasmids, generously provided by Dr. Angela Vincent, Dr. David Beeson, and Dr. Patrick Waters (University of Oxford, Oxford, UK), using branched polyethylenimine (Sigma, 408727). The culture medium was replenished the following day, and cells were harvested for CBA the day after.

To detect AChR-specific IgG binding capacity, AChR-expressing cells were preincubated with a 1:20 dilution of a serum sample or 1 µg/mL mAb for 1 hour at 4 °C. The cells were then stained with a rabbit anti-human IgG-Fcγ Alexa-Fluor™-647-conjugated secondary antibody (Jackson ImmunoResearch, 309-605-008). To detect AChR-IgA and IgM binding capacity, cells were preincubated with a 1:50 dilution of a serum sample or 1 µg/mL mAb for 1 hour at 4 °C, followed by staining with goat anti-human serum IgA-α chain specific-PerCP-conjugated (Jackson ImmunoResearch, 109-125-011) or goat anti-human IgM-Fc5µ-Alexa-Fluor™-647-conjugated secondary antibody (Jackson ImmunoResearch, 309-605-008), respectively. To detect AChR-IgG subclass binding capacity, AChR-expressing cells were preincubated with a 1:10 dilution of serum for 1 hour at 4 °C. The cells were then stained with either the mouse anti-human IgG1-hinge-AF647 (SouthernBiotech, 4E3), mouse anti-human IgG2-Fc-AF647 (SouthernBiotech, HP6002), mouse anti-human IgG3-hinge-AF647 (SouthernBiotech, HP6050), or mouse anti-human IgG4 pFc'-AF647 (SouthernBiotech, HP6023) secondary antibodies.

AChR-specific antibody binding on cells was determined by gating on AChR-rapsyn+ IgG+ cells using an LSR Fortessa (BD Biosciences). Flow cytometric data was analyzed using FlowJo software (v10.10.0). AChR-specific autoantibody binding strength was quantified by calculating the normalized mean fluorescence intensity (ΔMFI) as follows: $\Delta MFI = MFI_{AChR\text{-}GFP\text{-}positive} – MFI_{AChR\text{-}GFP\text{-}negative}$. A negative control mAb-58 IgG1 (AQP4-specific)[8] and positive controls mAb-637 IgG1, mAb-637 IgG2, mAb-637 IgG3, and mAb-637 IgG4, mAb-637 IgA, and mAb-637 IgM (all AChR-specific) were included in each assay.

**Complement assays**

AChR-specific antibody capacity to activate complement and deposit membrane attack complex (MAC) on the cell surface was assessed using triple-knockout HEK293T cells devoid of complement regulator genes CD46, CD55, and CD59 as previously described[9]. Briefly, cells were preincubated with a 1:20 dilution of a serum sample or 1 µg/mL mAb in the presence of normal human serum (NHS) as the consistent source of complement (25% final volume; Complement Technologies–Tyler, TX) for 3 hours at 37 °C. All serum samples were heat-inactivated (HI) to eliminate the influence of endogenous complement proteins. Cells were then fixed in 2% paraformaldehyde for 15 min at 4 °C. MAC deposition was detected by primary staining using a mouse anti-C9 neoantigen IgG2a antibody (Hycult Biotech, HM2167), followed by secondary antibody staining using a rat anti-mouse IgG2a-APC-conjugated (BioLegend, 407109).

Complement activity was additionally measured using C3d detection as a complementary approach. Briefly, HEK293T cells were preincubated with mAbs or serum samples in the presence of C8-depleted NHS for 1 hour at 37 °C (Complement Technologies–Tyler, TX). C3d deposition was detected by primary staining using a mouse anti-human C3d IgG1 antibody[10], generously provided by Dr. Joshua M. Thurman (University of Colorado, Anschutz Medical Campus, Aurora, Colorado, USA), Dr. Kelly Fahnoe, and Dr. Stefan Wawersik (Q32 Bio, Waltham, Massachusetts, USA), followed by secondary staining using a rat anti-mouse IgG1-PE-Cy7-conjugated antibody (RMG1-1, BioLegend). Complement activity was quantified by calculating the normalized mean fluorescence intensity (ΔMFI) as follows: $\Delta MFI = MFI_{AChR\text{-}GFP\text{-}positive} - MFI_{AChR\text{-}GFP\text{-}negative}$. A negative control mAb-58 IgG1 (AQP4-specific)[8] and a positive control mAb-637 IgG1 (AChR-specific) were included in each assay.

**ACh binding site blocking assay**

The capacity of autoantibodies to block α-bungarotoxin (α-BTX, serving as a proxy for ACh) binding to AChR was evaluated based on an approach that was previously described[11], with modifications that we reported[3] AChR-expressing HEK293T cells were preincubated with a 1:10 dilution of serum or 1 µg/mL mAb for 1 hour at 37 °C. The cells were then incubated with 1 µg/mL Alexa-Fluor™-647-conjugated α-BTX (Invitrogen, B35450). AChR-specific antibody blocking capacity was measured by the reduction in normalized MFI of Alexa-Fluor™-647-conjugated α-BTX, calculated as % Normalized $\Delta MFI_{\alpha\text{-}BTX} = (MFI_{observed} - MFI_{unstained}) / (MFI_{\alpha\text{-}BTX\text{-}treated} - MFI_{unstained}) \times 100$, where α-BTX-treated staining is set as the upper limit (MFI=100%) and unstained condition is set as the lower limit (MFI=0). An AChR-specific positive control mAb-01b, previously shown to block the ACh binding site[3], was included in each assay.

**AChR internalization assay**

The capacity of AChR-specific autoantibodies to induce internalization (modulation) of surface AChR was assessed as previously described[12] using the rhabdomyosarcoma CN21 muscle-type cell line. Briefly, CN21 cells were preincubated with a 1:50 dilution of serum or 1 µg/mL mAb in DMEM culture medium supplemented with 10% HI FBS for 16 hours at 37 °C to induce AChR internalization. To detect residual surface AChR, CN21 cells were then incubated with mouse anti-human AChR mcAb-3[13], generously provided by Dr. Vanda A. Lennon and Dr. John R. Mills (Mayo Clinic, Rochester, Minnesota, USA). The cells were then stained using an anti-mouse IgG1-PE-Cy7-conjugated antibody (BioLegend, 406614).

The capacity of AChR-specific antibodies to induce internalization of surface AChR was alternatively assessed using Zenon™ pHrodo™ iFL IgG Labeling Reagent. Briefly, CN21 cells were preincubated with a 1:50

dilution of serum or 1 µg/mL mAb for 1 hour at 37 °C. The cells were then incubated with 30 nM Zenon™ pHrodo™ iFL IgG Labeling Reagent for 1 hour at RT to label the Fc region of AChR-bound IgG. The cells were then incubated for 16 hours at 37 °C to allow the autoantibodies to induce AChR internalization. The fluorescent signal of the pHrodo labeling reagent was subsequently measured using a flow cytometer and also imaged using a fluorescence microscope.

The capacity of an autoantibody to internalize AChR was measured, in both assay formats, by the reduction in surface AChR, quantified by a scaled MFI, calculated as % Normalized MFI = $(MFI_{observed} - MFI_{unstained}) / (MFI_{untreated} - MFI_{unstained}) \times 100$, where the untreated (no test antibody) condition is set as the upper limit (MFI=100%), and the unstained (no primary and secondary antibodies) condition is set as the lower limit (MFI= 0). Negative controls included mAb-58 IgG1(AQP4-specific) and mAb-03 IgG1[3](only when the internalization was measured using mcAb-3). The mAb-03 IgG1 is AChR-specific but does not mediate the internalization of the AChR. The AChR-specific mAb-637 IgG1— previously shown to mediated internalization of the AChR—served as a positive control[3].

**Statistical analysis**

Statistical analyses were performed using GraphPad Prism software (version 10.2.1) and R (version 4.3.2). The positive detection threshold for the CBAs was determined by applying a cutoff value, calculated as the mean ΔMFI + 3×SD of healthy donors (HDs) in binding and complement assays, or the mean % Normalized MFI - 3SD of HDs in blocking and internalization assays. When comparing multiple groups, the Kruskal-Wallis test with Dunn's correction was used unless stated otherwise. Longitudinal analysis was performed using a longitudinal mixed model analysis in SAS (version 9.4). Correlation analyses were performed using Spearman's rank correlation test. A significance threshold of $P < 0.05$ was used and shown on plots as follows: *$P < 0.05$; ** $P < 0.01$, *** $P < 0.001$, and **** $P < 0.0001$. All the CBAs were performed in experimental triplicates for each sample. Samples were tested together in the same assay sessions to avoid batch effects.

**Supplementary Figures**

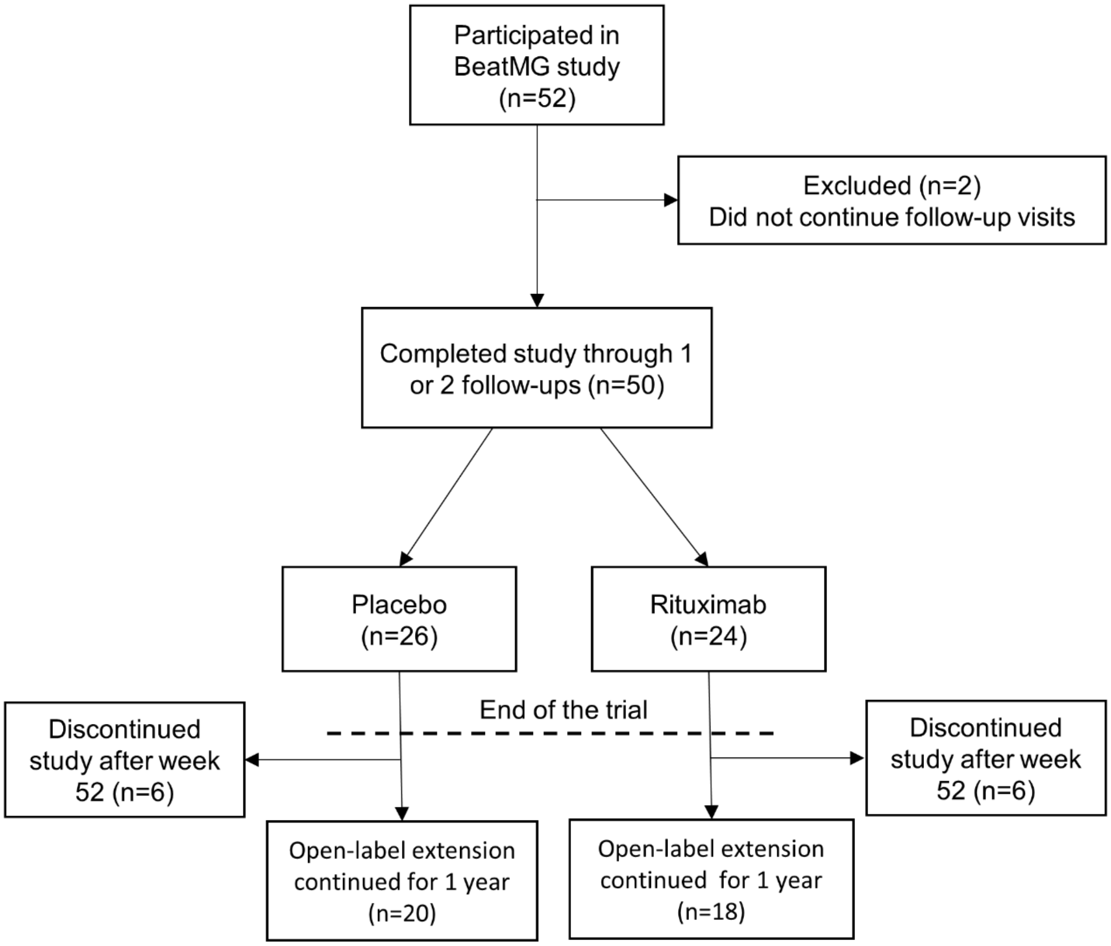


**Supplementary Figure 1. CONSORT flow diagram.**

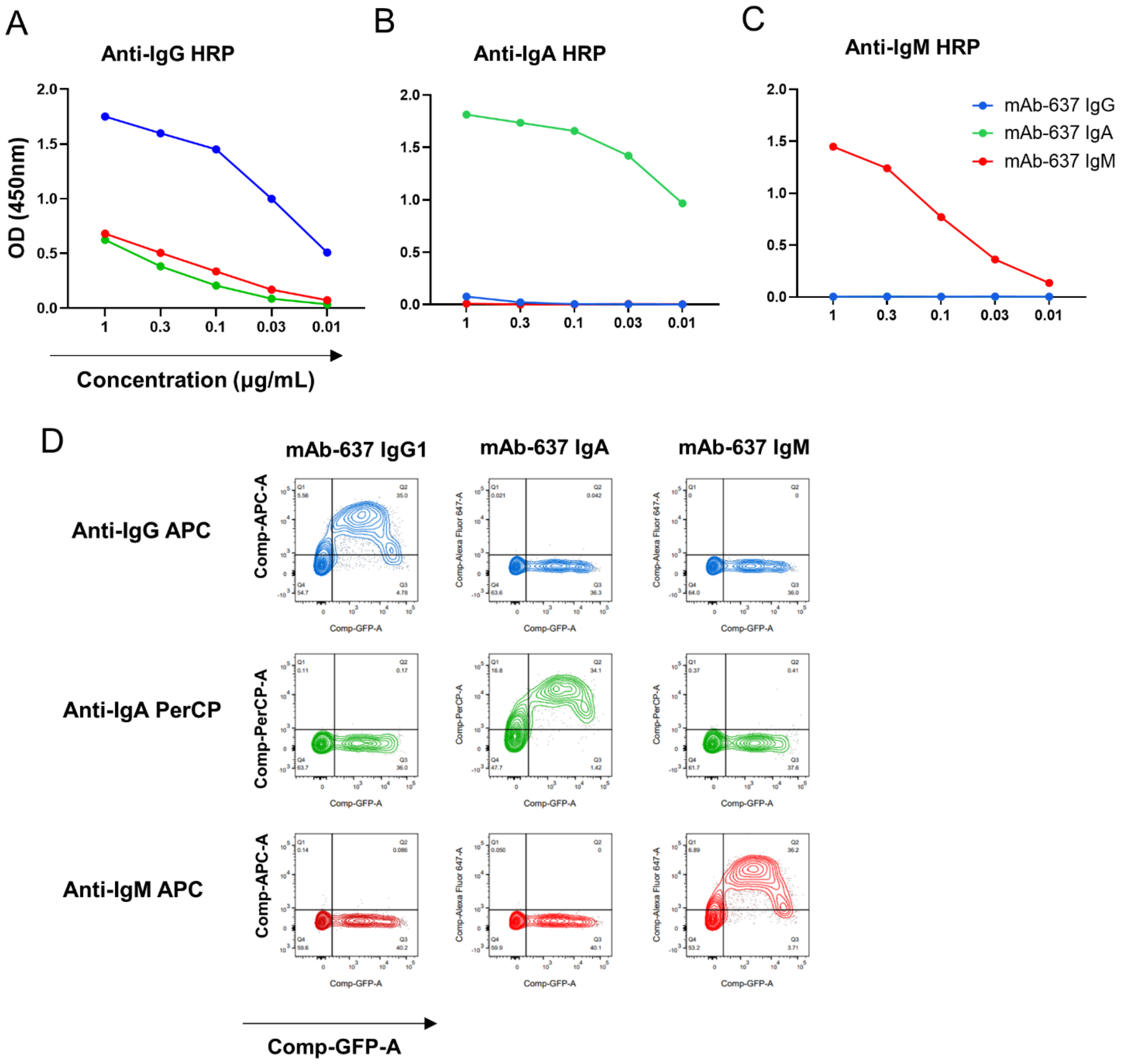


**Supplementary Figure 2. AChR-specific mAb isotype expression.** Confirmation of AChR-specific mAb isotype (IgG, IgM and IgA) expression and validation of commercial human Ig isotype secondary antibodies were performed. The AChR-specific recombinant mAb-637 was expressed as an IgG, IgA, or IgM, both ELISA and flow cytometry were used to test their expression and specificity of the commercial secondary antibodies. ELISA plates were coated with goat anti-human IgA+IgG+IgM at a concentration of 5 µg/mL, followed by incubation with serial dilutions of AChR mAb-637 IgG, IgA and IgM, ranging from 0.01 to 1 µg/mL. Recombinant mAb binding was detected using HRP-conjugated secondary antibodies. Titration plots showing the binding curves for (**A**) mAb-637 IgG, (**B**) IgA, and (**C**) IgM using HRP-conjugated isotype-specific secondary antibodies. (**D**) Contour plots, derived from FACS analysis, showing mAb-637 IgG, IgA, and IgM binding to AChR-expressing cells, detected with fluorophore-conjugated anti-human IgG, IgA, or IgM. Each data point (**A-C**) represents the mean of experimental duplicates.

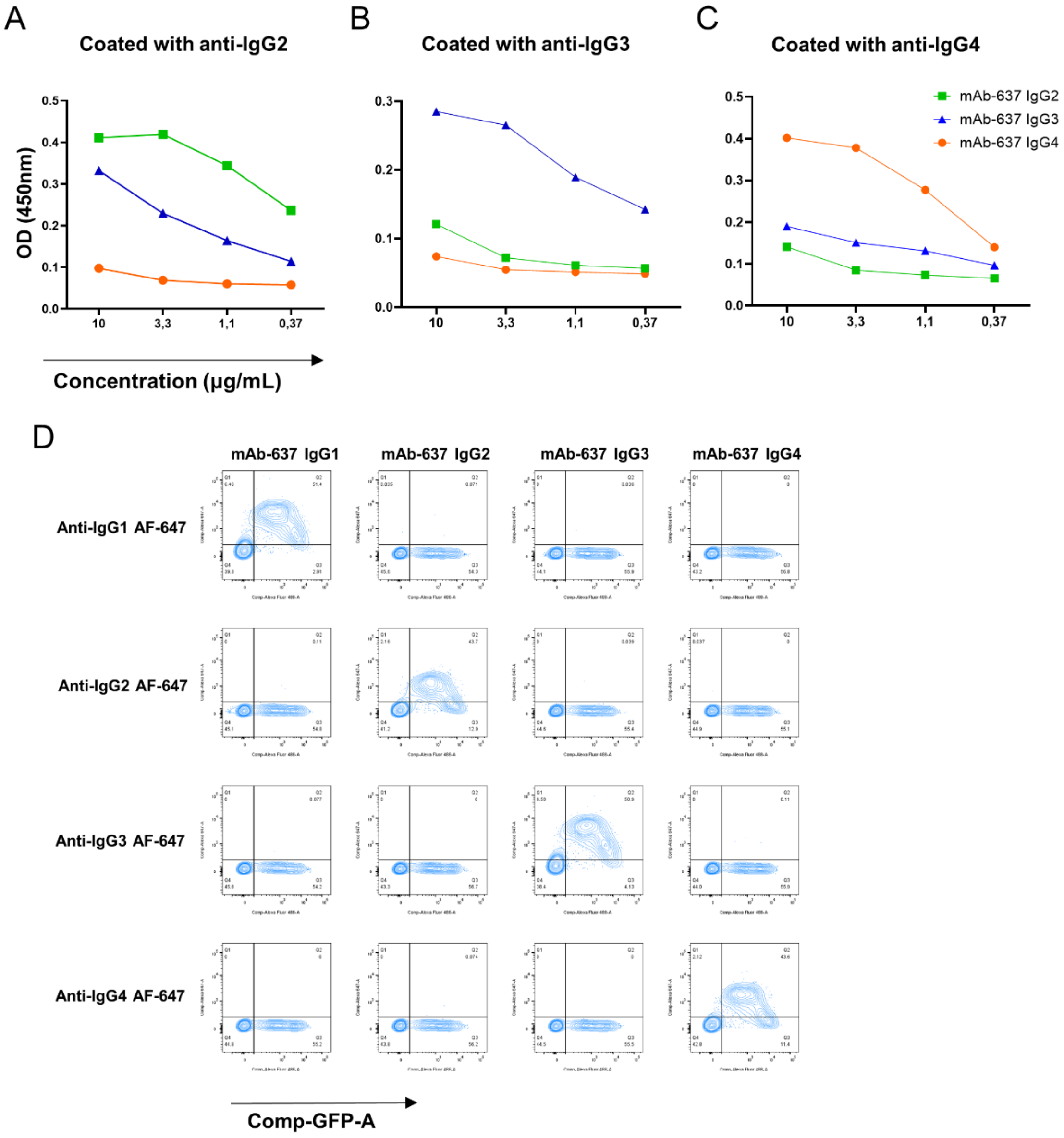


**Supplementary Figure 3. AChR-specific mAb IgG subclass expression.** Confirmation of AChR-specific mAb IgG subclass (IgG1, IgG2, IgG3, and IgG4) expression and validation of commercial human IgG subclass secondary antibodies were performed. The AChR-specific recombinant mAb-637 was expressed as an IgG1, IgG2, IgG3, or IgG4, both ELISA and flow cytometry were used to test their expression and specificity of the commercial secondary antibodies. ELISA plates were coated with subclass-specific antibodies for human IgG2, IgG3, and IgG4 at a concentration of 5 µg/mL, followed by incubation with serial dilutions of mAbs ranging from 0.037 to 10 µg/mL. Recombinant mAb binding was detected using HRP-conjugated secondary antibodies. Titration plots showing the binding curves for (**A**) mAb-637 IgG2, (**B**) IgG3, and (**C**) IgG4 using HRP-conjugated subclass-specific secondary antibodies. (**D**) Contour plots, derived from FACS analysis, showing mAb-637 IgG1, IgG2, IgG3, and IgG4 binding to AChR-expressing cells, detected with fluorophore-conjugated anti-human IgG1, IgG2, IgG3, and IgG4. Each data point (**A-C**) represents the mean of experimental duplicates.

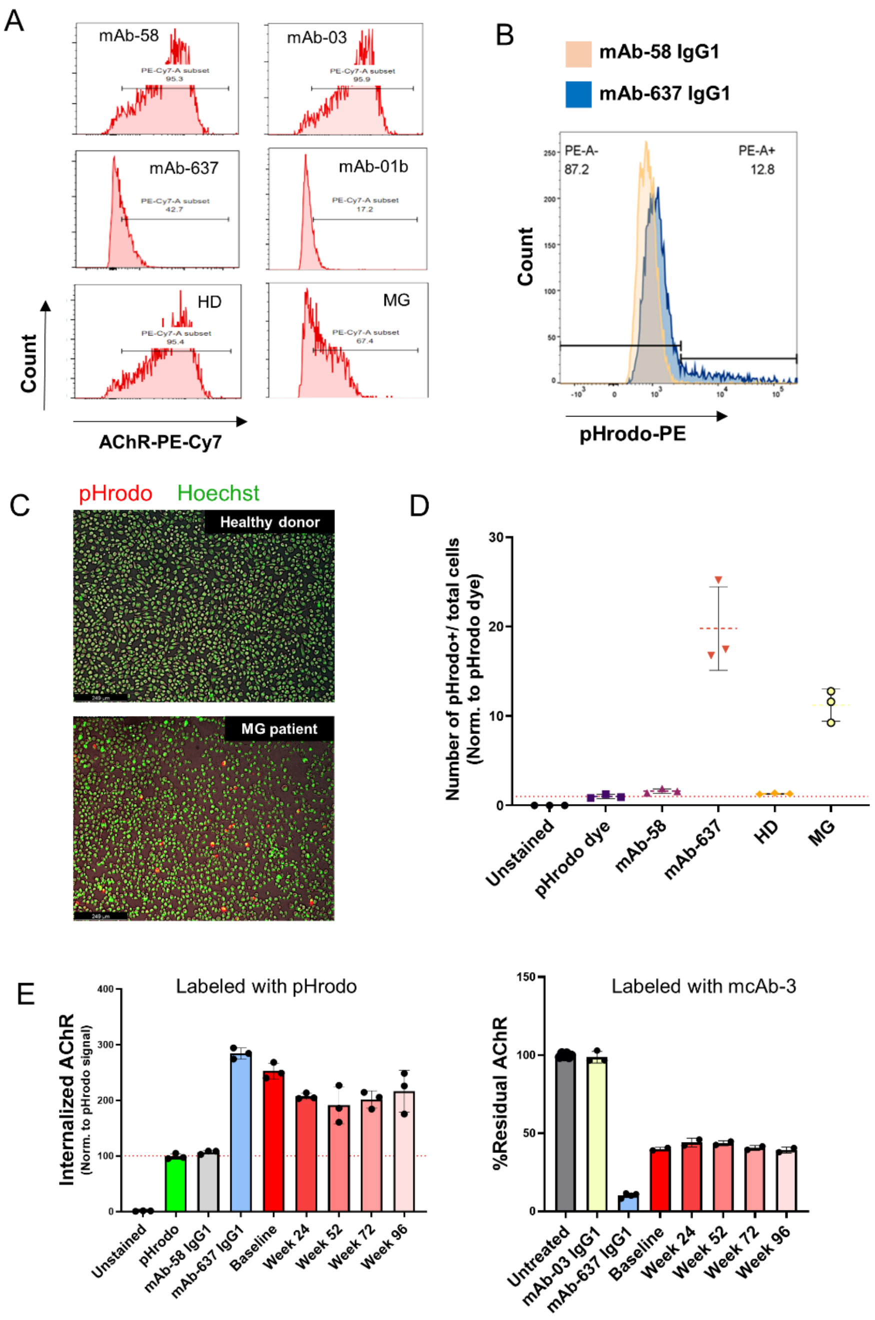


**Supplementary Figure 4. AChR antibody-mediated receptor internalization.** The capacity of AChR-specific autoantibodies to induce internalization (antigenic modulation) of surface AChR was assessed using the rhabdomyosarcoma CN21 cell line. Two independent assays were used, including an anti-AChR mAb (mcAb-3) and the Zenon™ pHrodo™ iFL IgG labeling reagent. To measure the residual AChR on the cell surface, cells were preincubated with a 1:50 dilution of serum or 1 µg/mL mAb for 16 hours at 37 °C to induce AChR internalization. Cells were then incubated with mouse anti-human AChR mcAb-3 and stained using an anti-mouse IgG1-PE-Cy7-conjugated antibody. To detect internalized AChR, cells were preincubated with a 1:50 dilution of serum or 1 µg/mL mAb for 1 hour at 37 °C, followed by incubation with 30 nM Zenon™ pHrodo™ iFL IgG Labeling Reagent for 1 hour at RT to label the Fc region of AChR-bound IgG. The cells were then incubated for 16 hours at 37 °C to allow the autoantibodies to induce AChR internalization. Histograms (**A**) showing residual AChR on the CN21 cell surface after incubation with positive and negative control mAbs, MG, and healthy donor serum samples. Negative control mAbs included mAb-58 IgG1 (AQP4-specific) and mAb-03 (AChR-specific); positive control mAbs included mAb-01b and mAb-637 (AChR-specific). Histograms (**B**) showing pHrodo fluorescent signal after internalization of AChR bound to the labeled mAb-637 IgG. Fluorescence microscopy image (**C**) showing internalized AChR. (**D**) Quantified AChR internalization (pHrodo+ cells/total cells) normalized to cells incubated only with pHrodo labeling reagent (no mAb set to 1.0). AChR internalization in CN21 cells was induced by longitudinal serum samples of a MG patient using (**E**) pHrodo labeling reagent (left) and mouse anti-human mcAb-3 (right; no mAb set to 100). A negative control mAb-58 IgG1 (AQP4-specific) and a positive control mAb-637 IgG1 (AChR-specific) were included. Error bars represent the mean ± SD. The dotted line represents the normalized pHrodo signal.

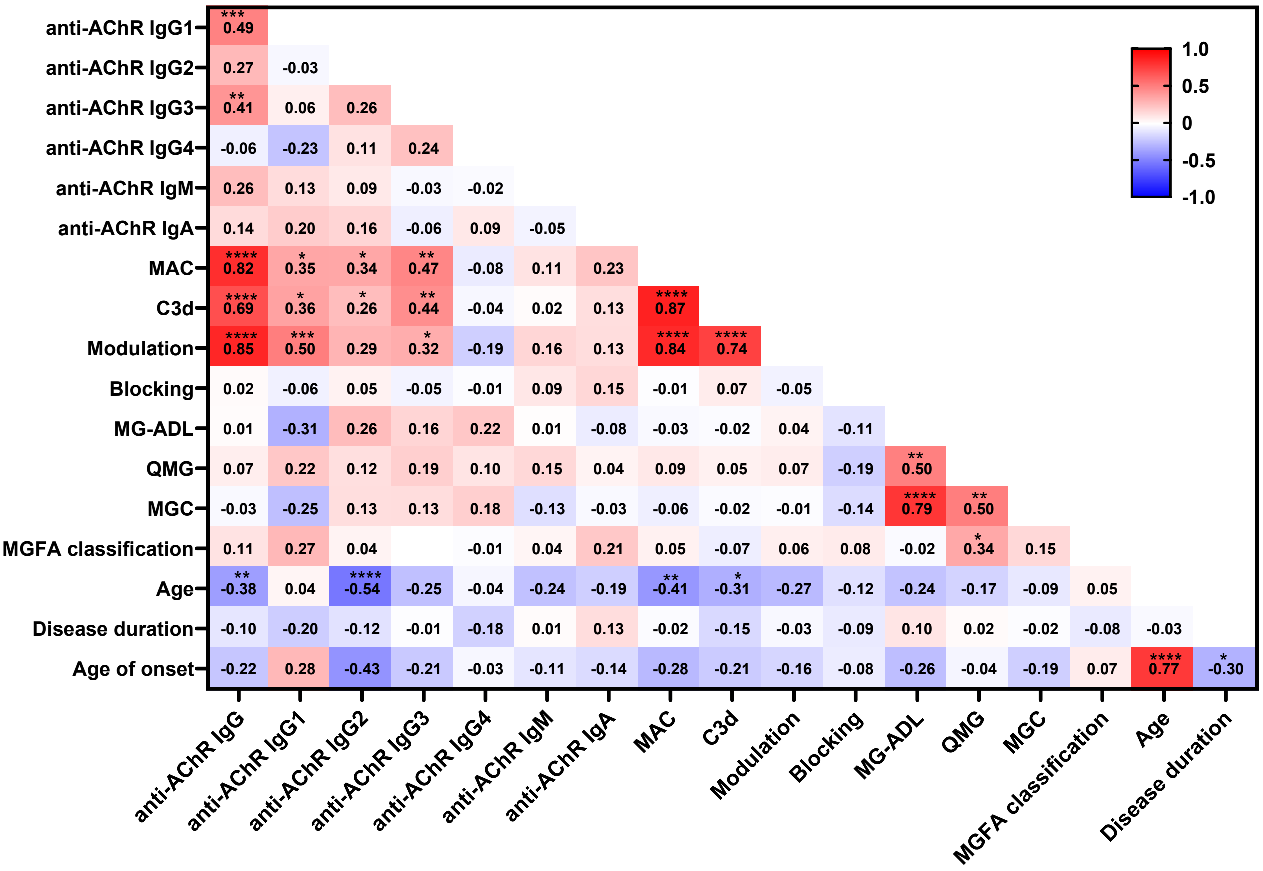


**Supplementary Figure 5. Association between serum AChR autoantibodies and clinical features.** The properties of AChR-specific serum autoantibodies and the pathogenic mechanisms they mediate were compared to key clinical features. Serum samples collected at baseline (N=46) were examined to investigate how autoantibody binding capacity and autoantibody-mediated pathogenic mechanisms, disease severity, MGFA classification, age, age of onset, and disease duration associate. Heatmap showing the Spearman correlation analysis, calculated using ΔMFI values for each individual patients, between AChR-specific autoantibodies binding and associated pathogenic mechanisms in the AChR patient cohort. The correlation coefficients (r) are shown on the heatmap. A significance threshold of $P < 0.05$ was used and is shown on plots when significance was reached: *$P < 0.05$; ** $P < 0.01$, *** $P < 0.001$, and **** $P < 0.0001$. The correlation coefficients (r) are shown on the heatmap. MG-ADL (MG-activities of daily living scale), QMG (quantitative myasthenia gravis), and MGC (myasthenia gravis composite).

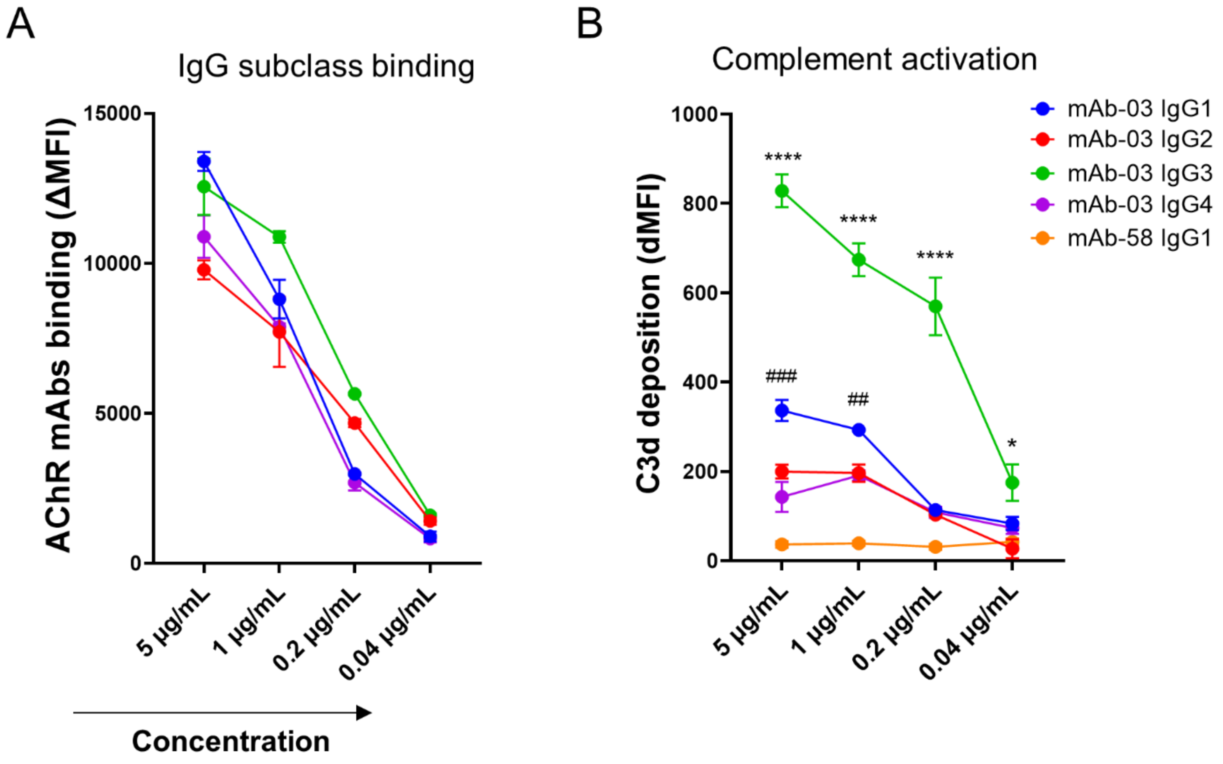


**Supplementary Figure 6. Activation of the classical complement pathway by AChR-specific IgM and IgG subclass antibodies.** Recombinant AChR-specific mAb-03 IgG subclasses were examined using an AChR-specific cell-based assay to evaluate their ability to bind AChR and activate the complement pathway. Titration plots showing (**A**) binding and (**B**) C3d deposition for mAb-03 IgG1, IgG2, IgG3, and IgG4 over a range of concentrations. A negative control mAb-58 IgG1 (AQP4-specific) was included. Each data point represents the mean of experimental triplicates, with error bars representing mean ± SD. Statistical analysis was performed using two-way ANOVA followed by Tukey's post-test. A significance threshold of $P < 0.05$ was used and is shown on plots when significance was reached: *$P < 0.05$; ** $P < 0.01$, *** $P < 0.001$, and **** $P < 0.0001$. Asterisks (*) compare mAb-03 IgG3 to IgG1; octothorpes (#) compare IgG1 to IgG2.

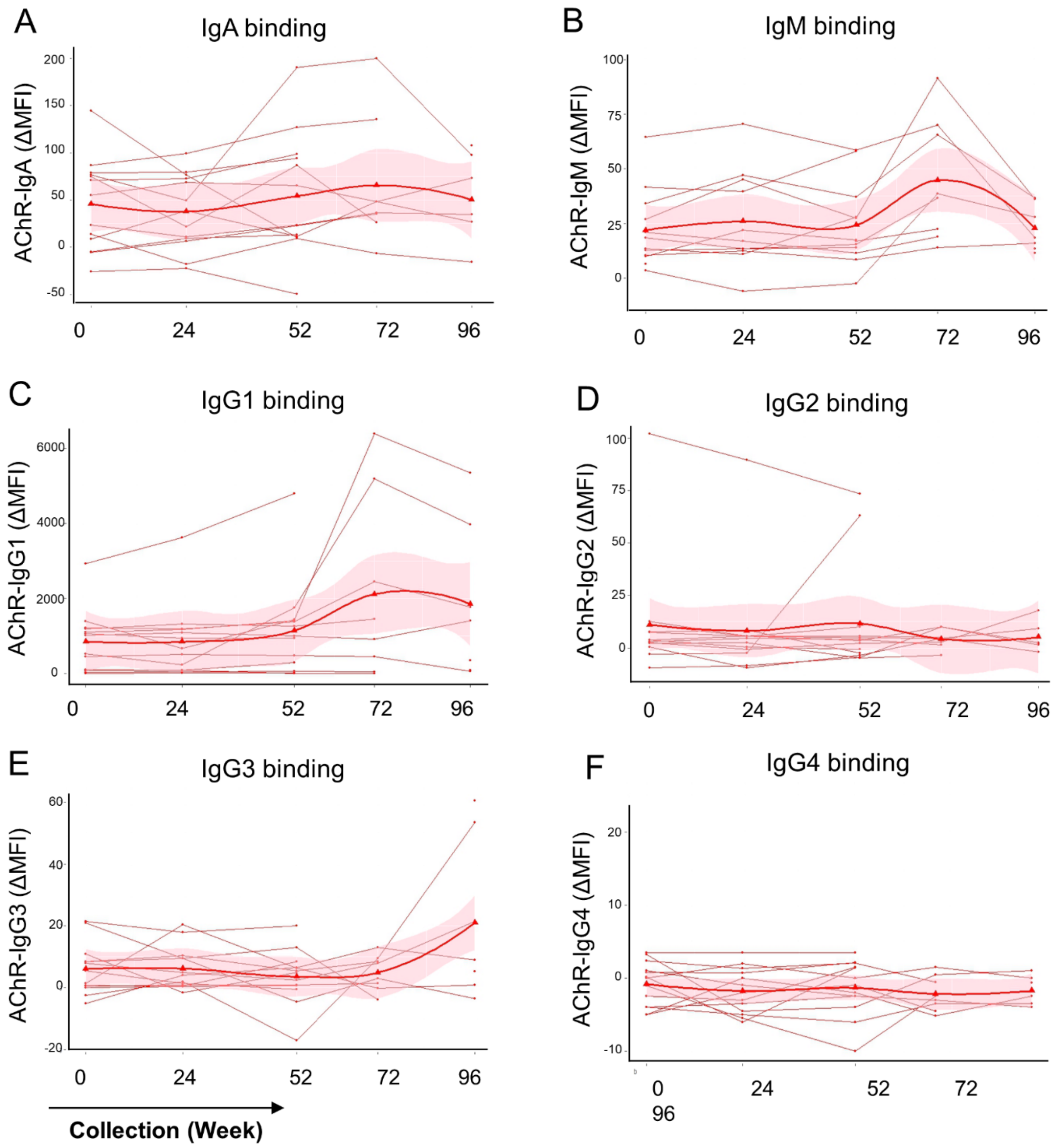


**Supplementary Figure 7. Temporal changes in AChR-IgM, IgA and IgG subclass binding capacity.** Serum samples collected longitudinally from a subset of patients in the placebo group (n=13) were examined using AChR-specific cell-based assays to determine the capacity of AChR-specific autoantibody binding within each patient over two years. Spaghetti plots show temporal changes in (**A**) AChR-IgA, (**B**) IgM, (**C**) IgG1, (**D**) IgG2, (**E**) IgG3, and (**F**) IgG4 binding to AChR-expressing cells measured in a cell-based assay. Each line represents a single patient (mean of experimental triplicates). Solid lines connect collection timepoints. The trend line (smooth bold curve) was created using the locally estimated scatterplot smoothing (LOESS) method. The shaded area on the graph depicts a 95% confidence interval (CI).

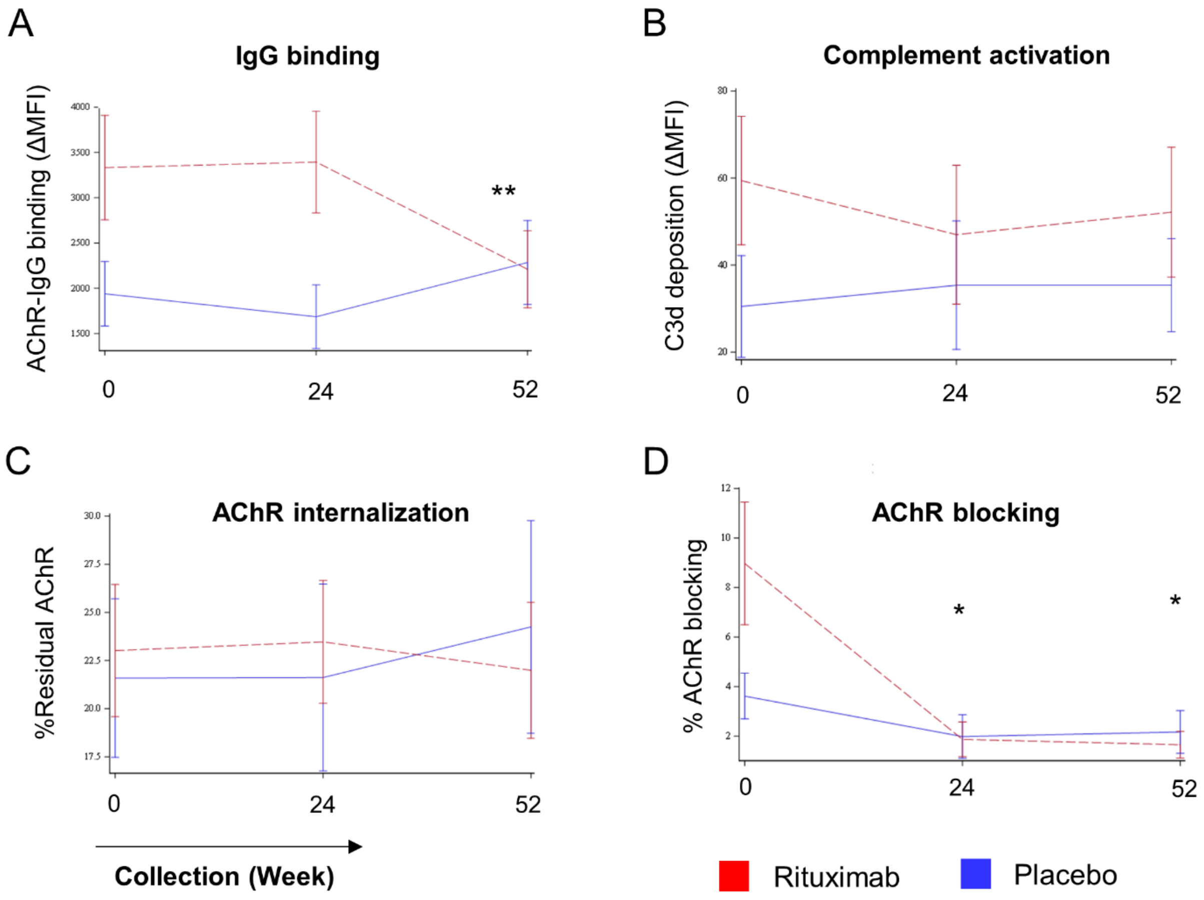


**Supplementary Figure 8. Effect of rituximab on the AChR-specific autoantibody repertoire and the associated pathogenic mechanisms.** Longitudinal serum samples from the placebo (n=26) and rituximab (n=24) groups were examined over one year using AChR-specific cell-based assays to evaluate the effects of rituximab on AChR-specific IgG and the associated pathogenic mechanisms. Graphs comparing (**A**) the mean AChR-IgG binding capacity between rituximab and placebo groups over time. (**B**) Graphs showing the magnitude of C3d deposition, (**C**) AChR internalization, (**D**) and blocking of the ACh binding site. Data were analyzed using a longitudinal mixed model. Error bars represent mean ± SEM, with assays performed in experimental triplicates. The AChR internalization and blocking percentages were calculated as (100 - (% residual AChR or % reduction of αBTX MFI)). Longitudinal statistical analysis compared accumulated changes between the rituximab and the placebo groups. A significance threshold of $P < 0.05$ was used and is shown on plots when significance was reached: *$P < 0.05$; ** $P < 0.01$.

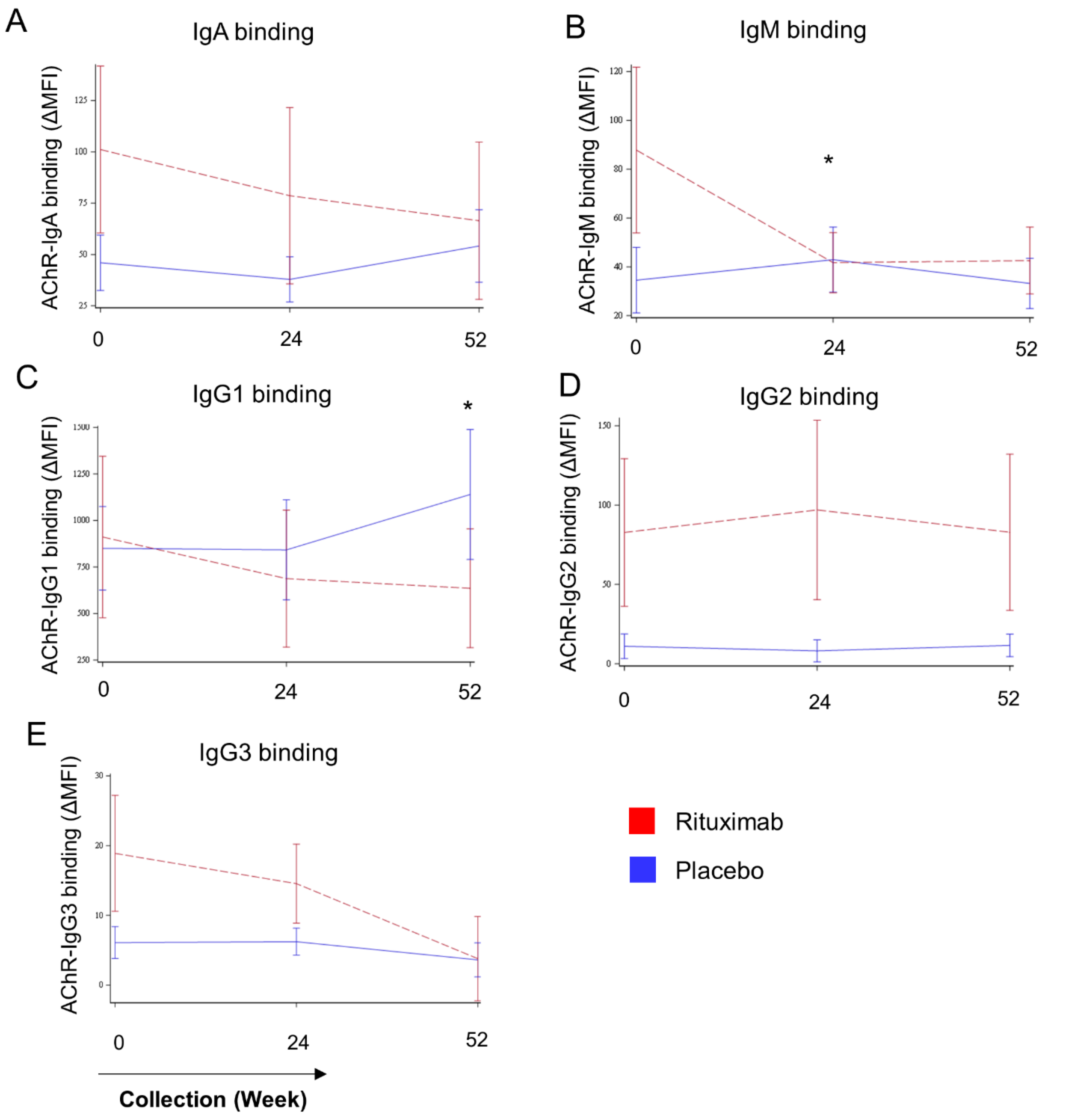


**Supplementary Figure 9. Effect of rituximab on AChR-specific autoantibody isotype and IgG subclass binding capacity.** Serum samples collected longitudinally from a subset of patients in the placebo (n=13) and rituximab (n=10) groups were examined over one year using AChR-specific cell-based assays to determine the effects of rituximab on AChR-specific autoantibody isotype and IgG subclasses. Graphs compare (**A**) the mean binding capacity for AChR-IgA, (**B**) AChR-IgM, (**C**) AChR-IgG1, (**D**) AChR-IgG2, and (**E**) AChR-IgG3 between rituximab and placebo groups over time. Data was analyzed using a longitudinal mixed model. Error bars represent mean ± SEM. Each line represents the mean for rituximab or placebo-treated groups, with assays performed in experimental triplicates. Longitudinal statistical analysis compared accumulated changes between the rituximab and the placebo groups. A significance threshold of $P < 0.05$ was used and is shown on plots when significance was reached: $^*P < 0.05$.

**Supplementary Table 1.** Study population characteristics recorded at baseline.

| **Clinical features and demographics** | **Total (n=50)** | **Rituximab (n=24)** | **Placebo (n=26)** |
|---|---|---|---|
| **Female** | 23 (46%) | 11 (45%) | 12 (46%) |
| **Age at enrollment, (y)** | 23-82 (54.7 ±17.2) | 23-81 (52.9 ±17.4) | 25-82 (56.5±16.8) |
| **Age of onset, (y)** | 20-81 (50.0 ±18.5) | 20-75 (45.5 ±19.4) | 21-81 (54.5 ±16.4) |
| **Disease duration, (y)** | 0-28 (5.2 ±5.7) | 0-28 (7.1 ±6.4) | 0-17 (3.4 ±4.0) |
| **Thymectomy** | 12 (24%) | 8 (33%) | 4 (15%) |
| **Relapse** | 15 (30%) | 5 (20%) | 10 (38%) |
| **Baseline clinical Disease activity** | | | |
| MGC score | 1-25 (9.5 ±5.13) | 1-25 (10.8 ±5.9) | 2-17 (8.3 ±3.9) |
| QMG score | 2-22 (9.9 ±4.5) | 2-22 (10.9 ±5.0) | 4-19 (9.0 ±3.8) |
| MG-ADL score | 0-12 (4.7 ±3.5) | 1-12 (5.6 ±3.5) | 0-12 (3.9 ±3.3) |
| **MGFA clinical class** | | | |
| Class I | 1 (2%) | 0 | 1 (3.8%) |
| Class II | 30 (60%) | 14 (58.3%) | 16 (61.5%) |
| Class III | 17 (34%) | 9 (37.5%) | 8 (30.7%) |
| Class IV | 2 (4%) | 1 (4.1%) | 1 (3.8%) |

Abbreviations: MGC = Myasthenia Gravis Composite; QMG = Quantitative Myasthenia Gravis; MG-ADL = Myasthenia Gravis-Activities of Daily Living; MGFA = Myasthenia Gravis Foundation of America. Values are displayed mean with standard deviation (SD) or n with frequency (%).
